# Inflammation-Related Biomarkers Linking Environmental Chemical Mixtures and Metabolic Dysfunction-Associated Steatotic Liver Disease: A Population-Based Study from NHANES 1999–2020

**DOI:** 10.64898/2026.10.06.26364867

**Authors:** Tingxian Guo, Yunhao Zhao, Shuang Wang, Chen Shen, Qi Jiang

## Abstract

**Objective:** Environmental pollutants are increasingly recognized as potential risk factors for metabolic dysfunction-associated steatotic liver disease (MASLD), yet the effects of mixed exposures and underlying mechanisms remain unclear. We evaluated associations between environmental chemical mixtures and MASLD and the mediating role of inflammation-related biomarkers.

**Methods:** We analyzed 60 environmental chemicals across six categories among 39,061 U.S. adults from the National Health and Nutrition Examination Survey (NHANES) 1999–2020. Principal Component Pursuit with Principal Component Analysis (PCP-PCA) identified latent exposure patterns. MASLD was identified using multisociety Delphi consensus criteria and the Hepatic Steatosis Index (HSI ≥ 36). Logistic regression, restricted cubic spline models, and mediation analysis were applied.

**Results:** PCP-PCA generated 5–17 dimensions per chemical category, with first-dimension variance contributions ranging from 30.70% to 98.88%. Six dimensions from four categories were retained, with phthalate esters (PAEs) and organophosphate flame retardants (OPFRs) showing the most consistent associations with MASLD. PAEs Dim.1, explaining 90.71% of variance, showed a significant non-linear dose-response association (P-nonlinear = 0.0001). Significant associations were identified for mono-isobutyl phthalate (MiBP; OR 1.12, 95% CI 1.07–1.19), mono(carboxyoctyl) phthalate (MCOP; OR 1.08, 95% CI 1.02–1.14), mono(carboxynonyl) phthalate (MCNP; OR 1.12, 95% CI 1.03–1.21), and bis(1,3-dichloro-2-propyl) phosphate (BDCP; OR 1.15, 95% CI 1.08–1.23). C-reactive protein (CRP) and red cell distribution width (RDW) were the primary mediators, accounting for 32.42% and 19.62% of the MiBP–MASLD association, respectively (both p < 0.0001).

**Conclusion:** Environmental co-exposure, particularly to PAEs and OPFRs, was associated with higher MASLD risk, with systemic inflammation potentially contributing to these associations.

## 1 Introduction

Metabolic dysfunction-associated steatotic liver disease (MASLD) is currently one of the most prevalent chronic liver diseases worldwide ^[1–5]^. Its incidence continues to rise and is closely associated with liver fibrosis ^[6]^, hepatocellular carcinoma ^[7]^, cardiovascular disease ^[8]^, and type 2 diabetes ^[9,10]^, posing a significant burden on public health systems ^[11]^. Although traditional risk factors such as genetic susceptibility, poor nutrition, and unhealthy lifestyles play important roles in the development of MASLD ^[12,13]^, increasing attention has recently been drawn to environmental pollution as a potential exogenous risk factor for MASLD ^[6,14]^.

Previous studies have suggested that various environmental chemical pollutants, including endocrine-disrupting chemicals, plasticizers, volatile organic compounds, flame retardants, and metals, may contribute to the development and progression of MASLD by interfering with hepatic lipid metabolism, inducing oxidative stress, and promoting inflammatory damage ^[14–17]^. However, most existing epidemiological evidence focuses on the association between exposure to individual pollutants and non-alcoholic fatty liver disease (NAFLD), the previous terminology that primarily relied on the exclusion of significant alcohol intake. By contrast, MASLD places greater emphasis on hepatic steatosis accompanied by cardiometabolic dysfunction. Comprehensive evaluations of the health effects of multiple pollutants in relation to MASLD holistically remain limited ^[18–21]^. In real-world settings, individuals are typically exposed to multiple low-dose pollutants simultaneously, and the resulting exposure profiles are often highly correlated and heterogeneous ^[22–26]^. Several mixture analysis methods have been developed in environmental epidemiology, such as Weighted Quantile Sum (WQS) regression and Bayesian Kernel Machine Regression (BKMR), which estimate overall mixture effects or identify interactions among exposures ^[22–26]^. However, these approaches are typically designed to estimate a single joint mixture effect from a moderate number of pre-selected chemicals, and may be limited when applied to high-dimensional exposure data spanning multiple chemical classes with distinct variance structures. In the present study, our primary aim was to identify underlying exposure patterns within each chemical category. We therefore applied Principal Component Pursuit (PCP) combined with Principal Component Analysis (PCA), which first decomposes the exposure matrix to separate systematic patterns from outliers and noise, and then extracts interpretable latent dimensions within each chemical category, enabling both dimension-level and individual chemical-level inference ^[27–29]^.

Mechanistically, the pathogenesis of MASLD is strongly influenced by chronic systemic inflammation ^[30]^. A growing body of research has demonstrated that various environmental pollutants, such as phthalates, organophosphate flame retardants, volatile organic compounds, and metals, can activate immune and inflammatory pathways, triggering systemic inflammatory responses that may disrupt hepatic metabolic homeostasis and contribute to tissue damage ^[30–32]^. In epidemiological studies, inflammation-related biomarkers have increasingly been investigated as potential biological mediators between environmental exposures and metabolic diseases ^[33–35]^. However, existing human evidence remains fragmented, with most studies focusing on individual pollutant classes, a limited number of inflammatory markers, or NAFLD rather than MASLD. Few population-based studies have systematically evaluated the mediating roles of multiple inflammation-related biomarkers in the associations between mixed environmental chemical exposures and MASLD. Such mechanistic evidence may contribute to causal inference and enhance understanding of the role of inflammation in underlying the associations between chemical pollutants and MASLD ^[15,36–39]^.

This study aimed to investigate the associations between mixed exposure of environmental chemicals and MASLD risk, and to elucidate the potential mediating role of inflammation-related biomarkers, thereby providing evidence for environmental risk prioritization and precision prevention of liver disease ^[6,15]^. Using nationally representative data from the U.S. National Health and Nutrition Examination Survey (NHANES) 1999–2020, we comprehensively evaluated 60 environmental chemicals across six major categories and examined both mixture-level and individual chemical associations with MASLD. We further assessed the potential mediating effects of multiple inflammation-related biomarkers to better understand the biological pathways linking environmental chemical exposures to MASLD.

## 2 Materials and methods

### 2.1 Data and study participants

NHANES is a population-based, cross-sectional study conducted by the Centers for Disease Control and Prevention (CDC) to evaluate the health and nutritional status of adults and children in the United States ^[40]^. NHANES surveys a nationally representative sample of approximately 5,000 people annually across various counties in the U.S. For this study, we utilized data from ten consecutive cycles of NHANES collected from 1999 to March 2020 Pre-pandemic (i.e., 1999-2000, 2001-2002, 2003-2004, 2005-2006, 2007-2008, 2009-2010, 2011-2012, 2013-2014, 2015-2016, and 2017-March 2020), focusing on participants with complete exposure and health outcome data relevant to the study objectives.

The initial sample included 107,622 NHANES participants from 1999-2020. Participants were excluded if they were under 20 years old, pregnant, had a history of cancer, lacked MASLD evaluation, or had missing exposure data. A total of 39,061 participants were included in the analysis (Figure 1A). The NHANES protocol was approved by the NCHS Institutional Review Board, and written informed consent was obtained from all study participants.

**Figure 1.**
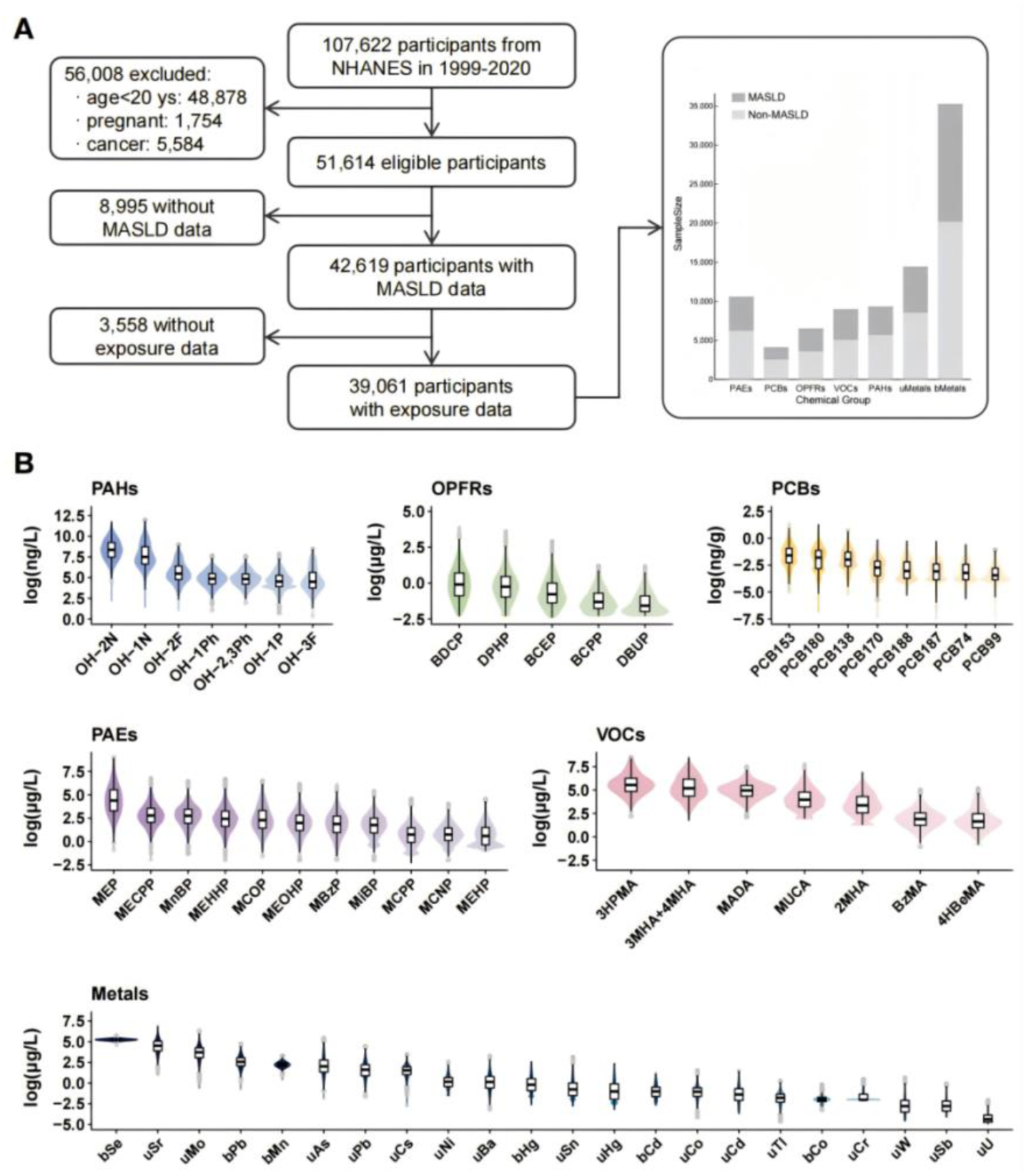
Research sample enrollment flow and concentration distributions of chemical pollutants. **A.** Flowchart of data enrollment for analysis. **B.** Distribution of exposure levels across six chemical categories. Abbreviations: PAEs, phthalate esters; PCBs, polychlorinated biphenyls; OPFRs, organophosphate flame retardants; VOCs, volatile organic compounds; PAHs, polycyclic aromatic hydrocarbons; uMetals, urinary metals; bMetals, blood metal; MASLD, Metabolic Dysfunction-Associated Steatotic Liver Disease

### 2.2 Measurement of exposure

This study included 60 environmental chemicals across six major categories: phthalate esters (PAEs), polychlorinated biphenyls (PCBs), organophosphate flame retardants (OPFRs), volatile organic compounds (VOCs), polycyclic aromatic hydrocarbons (PAHs), and metals. Whole blood and urine specimens were collected from participants at the time of enrollment and subsequently analyzed for pollutant or metabolite concentrations using high-performance liquid chromatography-electrospray ionization tandem mass spectrometry (HPLC-ESI-MS/MS) in CDC-certified laboratories following standardized protocols. Only chemicals with a detection rate exceeding 50% across the relevant NHANES cycles were included in the analysis. Urinary concentrations were measured for PAE metabolites (MCNP, MCOP, MCPP, MEP, MiBP, MnBP, MBzP, MEHP, MEHHP, MEOHP, and MECPP), OPFR metabolites (BCPP, BCEP, BDCP, DBUP, and DPHP), VOC metabolites (MUCA, 3HPMA, MADA, BzMA, 3MHA+4MHA, 2MHA, and 4HBeMA), PAH metabolites (OH-1N, OH-2N, OH-3F, OH-2F, OH-1Ph, OH-1P, and OH-2,3Ph), and 16 urinary metals (uAs, uCr, uHg, uBa, uCd, uCo, uCs, uMo, uPb, uSb, uSn, uTl, uW, uNi, uSr, and uU). Whole-blood concentrations were measured for PCBs (PCB74, PCB99, PCB138, PCB153, PCB170, PCB180, PCB187, and PCB188) and 6 blood metals (bCo, bPb, bCd, bHg, bSe, and bMn). Urinary analytes were normalized to creatinine concentration, and blood-based lipophilic compounds were adjusted using total lipids ^[40]^. Values below the limit of detection (LOD) were substituted with LOD divided by √2, a standard imputation approach widely adopted in environmental epidemiology for left-censored data ^[41]^.Detailed chemical names, abbreviations, measurement units, and cycle distributions are provided in Supplementary Table S1.

### 2.3 Assessment of MASLD

MASLD was defined based on the multisociety Delphi consensus statement ^[1]^. According to this definition, MASLD is diagnosed when hepatic steatosis is present in the absence of heavy alcohol use and viral hepatitis, accompanied by at least one of five cardiometabolic risk factors (CMRF): (1) body mass index (BMI) ≥ 25 kg/m², with a waist circumference ≥ 94 cm in men or ≥ 80 cm in women; (2) fasting blood glucose ≥ 100 mg/dL or hemoglobin A1c ≥ 5.7%, prior diagnosis of type 2 diabetes, or current anti-diabetic treatment; (3) blood pressure ≥ 130/85 mmHg or use of antihypertensive medication; (4) triglyceride levels ≥ 1.70 mmol/L or current lipid-lowering therapy; (5) HDL cholesterol<1.0 mmol/L in men or<1.3 mmol/L in women. Heavy alcohol use was defined as more than four drinks per day for men or more than three drinks per day for women ^[42–44]^. Viral hepatitis was identified based on serum HBsAg positivity, anti-HCV or HCV RNA positivity, or self-reported HBV/HCV infection.

Due to the absence of liver transient elastography data in NHANES cycles from 2003 to 2020, we introduced the Hepatic Steatosis Index (HSI) as a surrogate non-invasive indicator to assess hepatic fat accumulation based on liver enzymes (alanine aminotransferase (ALT) and aspartate aminotransferase (AST) and BMI). HSI has been widely validated and applied in epidemiological studies. It was calculated using the following published formula ^[24]^:

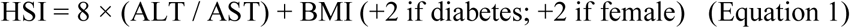

An additional 2 points were added for individuals with diabetes or for females. An HSI score of ≥36 was used as the threshold for hepatic steatosis. Participants were ultimately classified into MASLD and non-MASLD groups based on HSI score, metabolic abnormalities, and absence of heavy alcohol use or viral hepatitis.

### 2.4 Assessment of inflammation-related biomarkers

To evaluate the potential mediating role of inflammation in the associations between environmental chemical exposures and MASLD risk, we included 15 inflammation-related biomarkers as candidate mediators. These included one liver enzyme, gamma-glutamyl transferase (GGT), one acute-phase protein: C-reactive protein (CRP); one general inflammatory cell marker: white blood cell count (WBC); one hematological marker, red cell distribution width (RDW); and eleven composite inflammatory indices, including systemic immune-inflammation index (SII), neutrophil-to-lymphocyte ratio (NLR), platelet-to-lymphocyte ratio (PLR), systemic inflammation response index (SIRI), neutrophil-monocyte-to-lymphocyte ratio (NMLR), monocyte-to-lymphocyte ratio (MLR), derived NLR (dNLR), eosinophil-to-lymphocyte ratio (ELR), neutrophil-percentage-to-albumin ratio (NPAR), red cell distribution width-to-platelet ratio (RPR), and aggregate index of systemic inflammation (AISI).

GGT, CRP, WBC and RDW were directly obtained from the NHANES laboratory data. The remaining indices were derived from complete blood counts and other hematological parameters using established formulas, which are provided in the Supplementary Table S2. Collectively, these biomarkers reflect systemic inflammation, immune activation, and related hematological responses and have been widely used in epidemiological studies of chronic metabolic diseases. All 15 biomarkers were included in the mediation analyses to evaluate their potential indirect effects in the associations between chemical exposures and MASLD.

### 2.5 Principal component pursuit

To address high dimensionality and sparse outliers in the 60 environmental chemicals, we applied Principal Component Pursuit (PCP) before principal component analysis (PCA). PCP is a robust matrix decomposition method that separates an observed data matrix into a low-rank component and a sparse component. Let *X* ∈ ℝ*^n^*^×*p*^ denote the natural log-transformed exposure matrix, where *n* is the number of participants and *p* = 60 is the number of environmental chemicals. PCP decomposes *X* as:

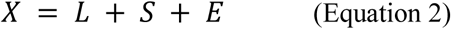

where *L* is a low-rank matrix representing coherent, shared exposure patterns across participants and chemicals; *S* is a sparse matrix capturing extreme observations or outliers; and *E* is a dense noise matrix. The decomposition was obtained by solving the following convex optimization problem:

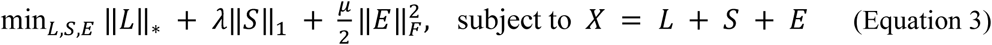

where ‖*L*‖_∗_ is the nuclear norm (sum of singular values), ‖*S*‖_1_ is the l_1_-norm (sum of absolute entries), and ‖*E*‖*_F_* is the Frobenius norm. *λ* > 0 controls the sparsity of *S*, and *μ* > 0 controls the residual noise. The optimization was solved using the alternating direction method of multipliers (ADMM; Boyd et al., 2011). The regularization parameter *λ* was set to 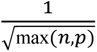 following Candès et al. (2011), and sensitivity analyses were performed using alternative *λ* values. After convergence, the low-rank component *L*, rather than the raw exposure matrix, was retained for subsequent PCA. Within each chemical category, PCP-PCA dimensions were retained until the cumulative variance explained reached at least 99%, thereby preserving the major exposure structure while excluding dimensions contributing only marginal residual variance. The resulting PCP-PCA scores were then used in the restricted cubic spline and multivariable logistic regression models described in Section 2.6. This PCP-PCA strategy has been shown to improve the stability and interpretability of exposure patterns in environmental epidemiology ^[27–29]^.

### 2.6 Statistical analysis

Baseline characteristics were summarized according to MASLD status. Continuous variables were presented as medians with interquartile ranges (IQRs), and categorical variables were presented as counts and percentages. The Shapiro-Wilk test was used to assess the normality of continuous variables. Because most environmental chemical concentrations were right-skewed, they were natural log-transformed before analysis. Differences between MASLD and non-MASLD groups were compared using Wilcoxon rank-sum tests for continuous variables and chi-square tests for categorical variables.

To reduce dimensionality and identify latent exposure patterns among the 60 environmental chemicals, we applied the PCP-PCA strategy described in Section 2.5. Following the extraction of major exposure dimensions, restricted cubic spline (RCS) models were used to evaluate potential non-linear dose-response relationships between the PCP-PCA scores and MASLD risk, with full adjustment for covariates. Multivariable logistic regression was then conducted to examine the associations between MASLD and individual chemicals that made major contributions to the MASLD-associated PCP-PCA dimensions, as indicated by component loadings. Three sequential models were constructed. Model 1 adjusted for urinary creatinine or total lipids, depending on the biological matrix of the chemical. Model 2 further adjusted for age, sex, race/ethnicity, country of birth, education level, family income-to-poverty ratio, and marital status. Model 3 additionally adjusted for smoking status, alcohol consumption, energy intake, and physical activity, and was considered the fully adjusted model.

Subsequently, mediation analysis was used to assess the indirect effects of inflammatory markers on the relationship between chemical exposure and MASLD, estimating total, direct, and indirect effects as well as mediation proportions. Non-parametric bootstrapping (n=1000) was applied to calculate 95% confidence intervals.

Covariates were selected based on prior literature and their relevance to both MASLD and environmental exposures. These included age, gender (male or female), race/ethnicity (Mexican American, Other Hispanic, Non-Hispanic, Other Race/Multiracial), marital status (married/living with partner, widowed/divorced/separated, and never married), country of birth (United States or other), education level (less than high school, high school graduate, and college or above), poverty-income ratio (<1.3, 1.3-3.5, and >3.5), smoking status (ever smoked or never smoked), alcohol consumption (light, moderate, or excessive), energy intake (continuous, in kcal/day), and physical activity (never, moderate, or vigorous). Urinary creatinine and total lipids were additionally adjusted for where appropriate, depending on the biological matrix of the chemical. All statistical analyses were conducted using R software (version 4.2.2). Statistical significance was defined as p<0.05. For analyses involving multiple comparisons, p-values were adjusted using the Benjamini-Hochberg false discovery rate (FDR) method.

## 3 Results

### 3.1 Characteristics of participants

The general characteristics of the study population are summarized in Table 1. The analysis included 39,061 participants aged 20 years and older, with an almost equal gender distribution (49.85% male and 50.15% female). The median age (IQR) was 48 (34-63) years for all participants, 43 (30-59) years for non-MASLD participants (N = 22,718), and 50 (37-63) years for those with MASLD (N = 16,343). Socioeconomic and lifestyle factors, including race/ethnicity, country of birth, marital status, smoking status, drinking status, energy intake, physical activity, and total lipids, showed significant differences between the MASLD and non-MASLD groups, as detailed in Table 1. Additionally, inflammation markers revealed a higher inflammatory burden in participants with MASLD, further highlighting the disparity between the two groups.

**Table 1.** Characteristics of the study population by MASLD status (N = 39,061).

| Characteristics | Total<br>(N = 39,061) | Non MASLD<br>(N = 22,718) | MASLD<br>(N = 16,343) | Statistic | P-Value |
| --- | --- | --- | --- | --- | --- |
| Gender, n (%) |  |  |  | 288.2386 | <0.0001 |
| Male | 19,471 (49.85) | 12,152 (53.49) | 7,319 (44.78) |  |  |
| Female | 19,590 (50.15) | 10,566 (46.51) | 9,024 (55.22) |  |  |
| Age, years | 48 (34-63) | 43 (30-59) | 50 (37-63) | 531.5289 | <0.0001 |
| Race/ethnicity, n (%) |  |  |  | 80.7689 | 0.0013 |
| Mexican American | 7,010 (18.02) | 4,015 (17.75) | 2,995 (18.39) |  |  |
| Other Hispanic | 3,043 (7.82) | 1,641 (7.25) | 1,393 (8.55) |  |  |
| Non-Hispanic | 24,651 (63.36) | 14,275 (63.11) | 10,376 (63.72) |  |  |
| Others | 4,209 (10.82) | 2,690 (11.89) | 1,519 (9.33) |  |  |
| Country of birth, n (%) |  |  |  | 10.3659 | 0.0013 |
| US | 28,051 (71.85) | 16,174 (71.23) | 11,877 (72.72) |  |  |
| Others | 10,988 (28.15) | 6,532 (28.77) | 4,456 (27.28) |  |  |
| Poverty Income Ratio, n (%) |  |  |  | 9.6470 | 0.0080 |
| <1.3 | 10,990 (30.98) | 6,521 (31.59) | 4,469 (30.13) |  |  |
| 1.3-3.5 | 13,430 (37.86) | 7,786 (37.72) | 5,644 (38.05) |  |  |
| >3.5 | 11,056 (31.16) | 6,337 (30.70) | 4,719 (31.82) |  |  |
| Education, n (%) |  |  |  | 9.9649 | 0.0069 |
| Less than High school | 9,947 (26.72) | 5,835 (26.87) | 4,112 (26.52) |  |  |
| High school | 19,341 (51.96) | 11,150 (51.34) | 8,191 (52.83) |  |  |
| College or above | 7,934 (21.32) | 4,734 (21.80) | 3,200 (20.64) |  |  |
| Marital, n (%) |  |  |  | 290.7403 | <0.0001 |
| Married/Living with partner | 21,847 (59.29) | 12,266 (56.98) | 9,581 (62.53) |  |  |
| Widowed/divorced/Separated | 7,996 (21.70) | 4,536 (21.07) | 3,460 (22.58) |  |  |
| Never married | 7,006 (19.01) | 4,725 (21.95) | 2,281 (14.89) |  |  |
| Smoking status, n (%) |  |  |  | 285.8041 | <0.0001 |
| Smoked | 17,505 (44.86) | 10,998 (48.47) | 6,507 (39.84) |  |  |
| Never smoked | 21,515 (55.14) | 11,691 (51.53) | 9,824 (60.16) |  |  |
| Drinking status, n (%) |  |  |  | 2,017.7401 | <0.0001 |
| Light alcohol consumption | 7,684 (21.41) | 3,420 (16.37) | 4,264 (28.43) |  |  |
| Moderate alcohol consumption | 10,970 (30.57) | 5,359 (25.65) | 5,611 (37.41) |  |  |
| Excessive alcohol consumption | 17,236 (48.02) | 12,111 (57.98) | 5,125 (34.17) |  |  |
| Energy Intake, kcal/d | 2,009 (1,474-2,705) | 2,072 (1,527-2,805) | 1,939 (1,413-2,596) | 98.9930 | <0.0001 |
| Activity, n (%) |  |  |  | 69.7110 | <0.0001 |
| Never | 11,698 (29.96) | 6,480 (28.53) | 5,218 (31.94) |  |  |
| Moderate | 15,537 (39.79) | 9,053 (39.87) | 6,484 (39.69) |  |  |
| Vigorous | 11,810 (30.25) | 7,176 (31.60) | 4,634 (31.60) |  |  |
| Family Size | 3 (2-4) | 3 (2-4) | 3 (2-4) | 1.2597 | 0.2617 |
| SleepTime, h | 7 (6-8) | 7 (6-8) | 7 (6-8) | 0.1324 | 0.7160 |
| Creatinine, µmol/L | 99.89 (58.34-148.51) | 97.24 (54.80-146.74) | 102.54 (61.88-149.40) | 1.4937 | 0.2216 |
| Total Lipids, µmol/L | 10.45 (9.02-11.95) | 10.26 (8.85-11.79) | 10.69 (9.30-12.10) | 100.0113 | <0.0001 |
| Inflammation |  |  |  |  |  |
| GGT, IU/L | 21 (14-31) | 19 (14-30) | 23 (16-34) | 691.8599 | <0.0001 |
| CRP (mg/L) | 0.30 (0.11-0.84) | 0.36 (0.10-1.30) | 0.45 (0.19-1.27) | 917.1172 | <0.0001 |
| WBC (1000 cells/uL) | 6.90 (5.70-8.30) | 6.80 (5.60-8.30) | 7.10 (5.90-8.50) | 217.3231 | <0.0001 |
| SIRI | 1.02 (0.69-1.49) | 1.02 (0.69-1.50) | 1.03 (0.70-1.49) | 12.5426 | 0.0004 |
| PLR | 120 (95-152) | 117.58 (93.16-148.39) | 118.18 (93.75-148.57) | 43.6450 | <0.0001 |
| NLR | 1.93 (1.44-2.55) | 1.91 (1.43-2.55) | 1.92 (1.44-2.50) | 2.4259 | 0.1193 |
| dNLR | 0.84 (0.80-0.87) | 0.84 (0.80-0.87) | 0.84 (0.80-0.87) | 11.1477 | 0.0008 |
| SII | 165.00 (107.42-312.63) | 209.84 (118.77-421.67) | 176.36 (112.60-342.98) | 135.0937 | <0.0001 |
| MLR | 0.25 (0.20-0.33) | 0.26 (0.20-0.33) | 0.25 (0.20-0.32) | 32.4197 | <0.0001 |
| NMLR | 2.19 (1.67-2.86) | 2.19 (1.67-2.89) | 2.18 (1.67-2.82) | 1.5453 | 0.2138 |
| AISI | 245.94 (158.44-380.68) | 240.35 (153.63-375.65) | 245.44 (165.70-388.05) | 39.7178 | <0.0001 |
| ELR | 1.92 (1.44-2.55) | 1.93 (1.44-2.58) | 1.92 (1.44-2.50) | 1.5242 | 0.2170 |
| NPAR | 13.64 (12.02-15.30) | 13.49 (11.88-15.15) | 13.83 (12.20-15.48) | 104.2726 | <0.0001 |
| RDW (%) | 13.10 (12.50-13.80) | 12.90 (12.40-13.70) | 13.20 (12.60-14.00) | 414.2884 | <0.0001 |
| RPR | 0.05 (0.05-0.06) | 0.05 (0.05-0.06) | 0.05 (0.05-0.06) | 4.1635 | 0.0413 |

$\chi^2$ statistics are reported for categorical variables; Kruskal-Wallis H statistics are reported for continuous variables. Abbreviations: MASLD, Metabolic Dysfunction-Associated Steatotic Liver Disease; CRP, C-reactive protein; WBC, white blood cell count; GGT, gamma-glutamyl transferase; PLR, platelet-to-lymphocyte ratio; NLR, neutrophil-to-lymphocyte ratio; dNLR, derived neutrophil/lymphocyte ratio; SII, systemic immune-inflammation index; MLR, monocyte-to-lymphocyte ratio; NMLR, neutrophil-monocyte to lymphocyte ratio; AISI, aggregate index of systemic inflammation; NPAR, neutrophil-percentage-to-albumin ratio; ELR, eosinophil-to-lymphocyte ratio; RDW, Red Cell Distribution Width; RPR, Red Cell Distribution Width to Platelet Count Ratio

### 3.2 Exposure of Environment Chemicals

A total of 60 chemicals across six categories were analyzed, including PAHs, OPFRs, PCBs, PAEs, VOCs, and metals. The analysis revealed substantial variation in concentrations. PAHs showed a wide concentration range, from 1.4 to 161,363 ng/L. PAEs and VOCs also showed broad concentration ranges, with PAEs ranging from 0.06 to 9,040.7 μg/L and VOCs from 0.35 to 7,090 μg/L. In contrast, OPFRs and PCBs had narrower observed ranges, with OPFRs ranging from 0.06 to 50.7 μg/L, and PCBs between 0 and 3.71 ng/g. Metals demonstrated a broad range from 0 to 1,007.81 μg/L, reflecting variability based on the specific element. Detailed exposure data, including LOD, GM, and IQR, are provided in Supplementary Table S1.

### 3.3 PCP-PCA Analysis of Chemical Categories

PCP-PCA was applied to characterize the variance structure and latent exposure patterns within the six chemical categories. Dimensions required to explain approximately 99% of the cumulative variance within each category were retained as the major exposure dimensions and are highlighted in purple in Figure 2A. For PAEs, VOCs, and PAHs, the cumulative variance exceeded 99% within the first two or three dimensions, indicating relatively concentrated exposure structures. In contrast, OPFRs, PCBs, and metals required a larger number of dimensions to capture a comparable proportion of the total variance, suggesting more dispersed exposure structures. Detailed eigenvalues, variance explained, and cumulative variance for all PCP-PCA dimensions are provided in Supplementary Table S3.

**Figure 2.**
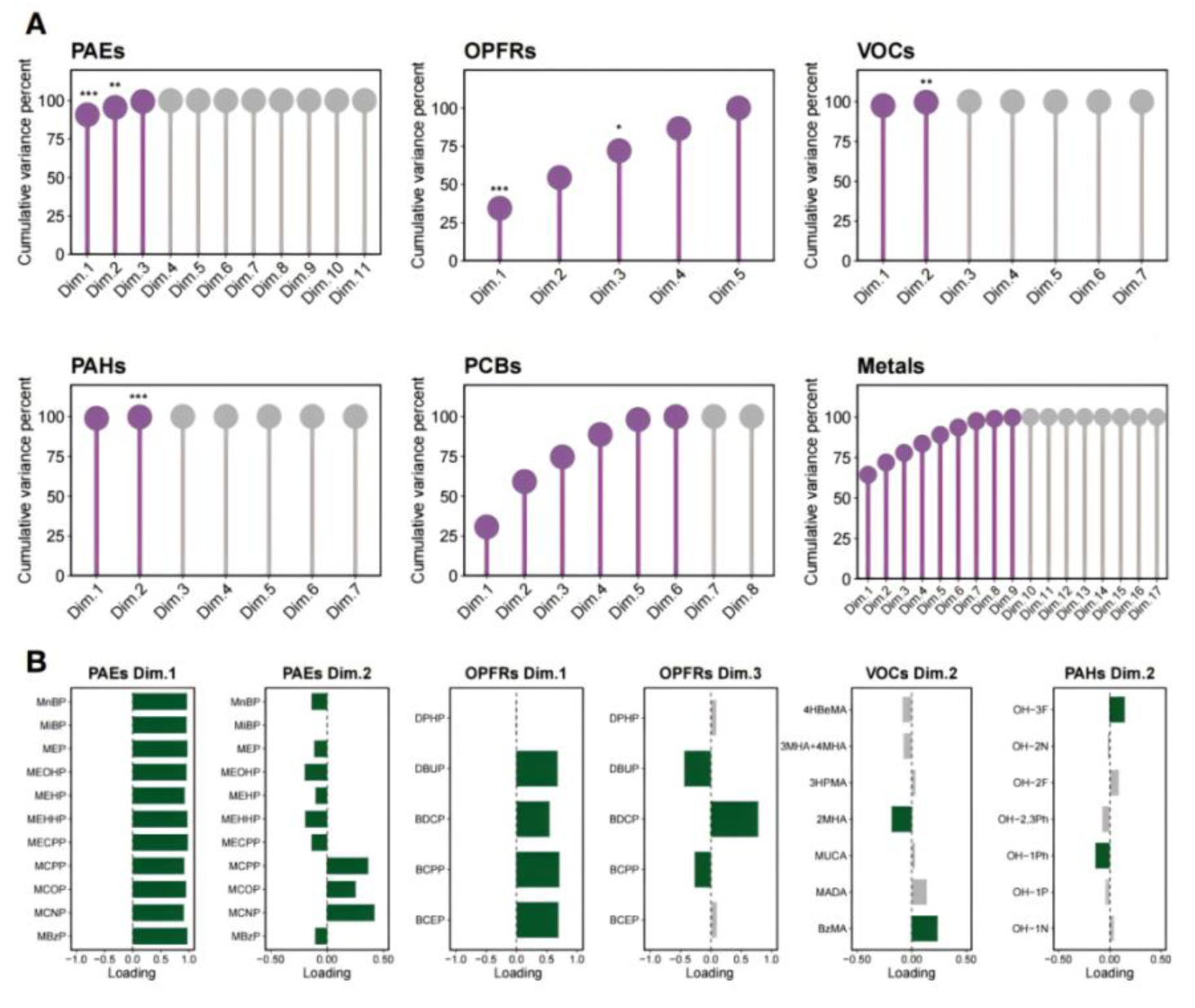
PCP-PCA-Derived Exposure Patterns Across Chemical Categories. **A.** Cumulative variance explained by PCP-PCA dimensions within each chemical category. **B.** Loadings and eigenvectors of the principal components significantly associated with MASLD for each chemical category. Purple markers indicate dimensions retained to represent the major exposure structure based on cumulative variance explained; gray markers indicate subsequent dimensions. Asterisks denote FDR-adjusted associations with MASLD among the retained dimensions. Abbreviations: PAEs, phthalate esters; PCBs, polychlorinated biphenyls; OPFRs, organophosphate flame retardants; VOCs, volatile organic compounds; PAHs, polycyclic aromatic hydrocarbons; MASLD, Metabolic Dysfunction-Associated Steatotic Liver Disease; Dim., dimension

For the six dimensions retained for downstream analyses, as described in Section 3.4, component loadings were examined to characterize the contributions of individual chemicals to each latent exposure pattern (Figure 2B). Chemicals with absolute loading values greater than 1/n, where n represents the number of chemicals within the corresponding category, were highlighted. For PAEs, most individual chemicals showed substantial loadings on Dim.1, whereas Dim.2 represented a more heterogeneous pattern of positive and negative loadings. For OPFRs, several chemicals contributed substantially to Dim.1 and Dim.3. In contrast, the selected VOC and PAH dimensions were characterized by substantial loadings from a smaller subset of individual chemicals. These loading patterns were subsequently used to guide the individual-chemical analyses.

### 3.4 Associations between major PCP-PCA dimensions and MASLD Risk

Associations between the major PCP-PCA dimensions and MASLD risk were evaluated with adjustment for multiple comparisons. Six dimensions from four chemical categories showed significant associations with MASLD after Benjamini-Hochberg FDR adjustment: PAEs Dim.1 (OR = 1.06, 95% CI: 1.04–1.08; FDR-adjusted P < 0.001), PAEs Dim.2 (OR = 1.15, 95% CI: 1.06–1.24; FDR-adjusted P = 0.005), OPFRs Dim.1 (OR = 0.87, 95% CI: 0.82–0.93; FDR-adjusted P < 0.001), OPFRs Dim.3 (OR = 0.88, 95% CI: 0.80–0.96; FDR-adjusted P = 0.030), VOCs Dim.2 (OR = 1.49, 95% CI: 1.18–1.89; FDR-adjusted P = 0.007), and PAHs Dim.2 (OR = 0.45, 95% CI: 0.32–0.62; FDR-adjusted P < 0.001). No significant associations were observed for the major dimensions of PCBs or metals after FDR adjustment. These six dimensions were carried forward for dose-response analyses and detailed interpretation of component loadings. Complete results for the candidate PCP-PCA dimensions are provided in Supplementary Table S4.

### 3.5 Dose-response analysis of PCP-PCA scores and MASLD risk

To further explore the significant dimensions identified in the PCP analysis, we employed RCS curves to examine the dose-response relationships between the PCA scores of these dimensions and the risk of MASLD (Figure 3). This approach was chosen to uncover potential nonlinear associations, providing a more nuanced understanding of the relationship between pollutant exposure and MASLD risk. A particularly notable result emerged from the analysis of PAEs Dim.1, which showed a significant overall association with MASLD risk (P-total<0.0001). The odds of MASLD generally increased with higher PCA scores, with evidence of non-linearity also observed (P-nonlinear=0.0001). This finding is especially important given that, in our previous PCP analysis, PAEs Dim.1 explained 90.71% of the variance in the overall chemical exposure profile. The dominance of this component suggests that PAEs Dim.1 is a major driver in the relationship between phthalate exposure and MASLD risk, making it a key focus for this research.

**Figure 3.**
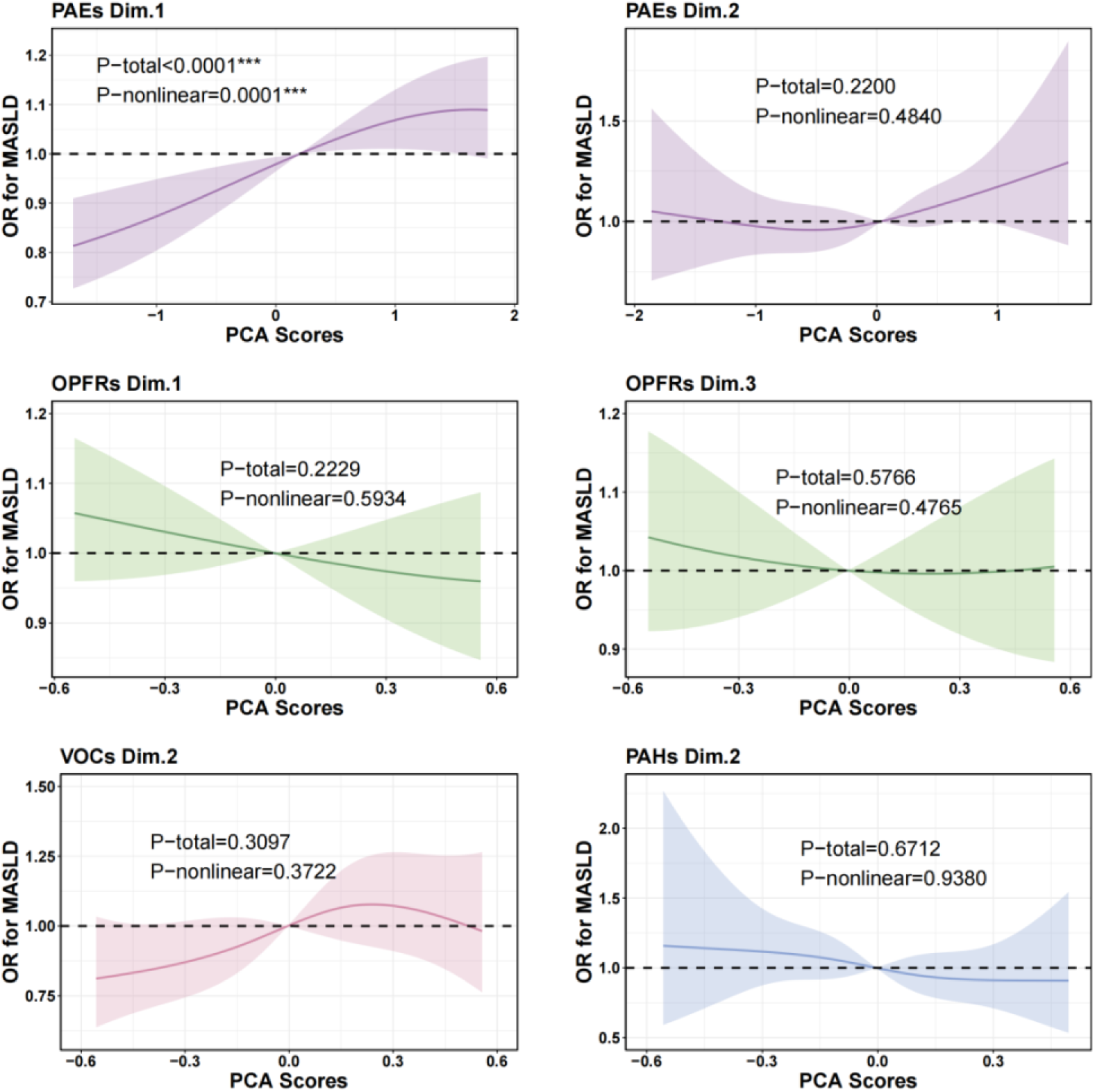
Dose-response relationship between PCA scores of chemical pollutants and MASLD. RCS curves for significant dimensions identified in the PCP analysis, depicting dose-response relationships between PCA scores and MASLD. The x-axis represents PCA scores, while the y-axis indicates OR for MASLD. The dark-colored solid line represents the OR values, and the light-colored shading around the line denotes the 95% CI. This model is fully adjusted according to Model 3 in the analysis of total effects. Abbreviations: PAEs, phthalate esters; OPFRs, organophosphate flame retardants; VOCs, volatile organic compounds; PAHs, polycyclic aromatic hydrocarbons; MASLD, Metabolic Dysfunction-Associated Steatotic Liver Disease

In addition, we observed a rising trend in the dose-response relationship for PAEs Dim.2 and VOCs Dim.2, even though these associations did not reach statistical significance. Specifically, as the PCA scores for these dimensions increased beyond a moderate level, MASLD risk showed an upward trajectory. PAEs Dim.2 and VOCs Dim.2 showed upward or curved patterns at higher PCA scores, but neither the overall nor non-linear associations reached statistical significance. Therefore, these patterns should be interpreted as exploratory and not as evidence of significant dose-response relationships. For OPFRs Dim.1, OPFRs Dim.3, and PAHs Dim.2, no clear dose-response relationship was observed.

### 3.6 Associations between major individual pollutants and MASLD risk

In this section, we further examined the associations between individual environmental pollutants and MASLD risk, as identified in the PCP analysis, using logistic regression. The detailed results are shown in Figure 4. The individual-level analysis was broadly consistent with the PCP-PCA and RCS findings, with significant associations mainly observed for PAEs and OPFRs. Among the individual chemicals in these categories, specific compounds, such as MiBP (in the PAEs category) and BDCP (in the OPFRs category), were strongly associated with MASLD. In contrast, VOCs and PAHs showed minimal or non-significant effects on MASLD risk.

**Figure 4.**
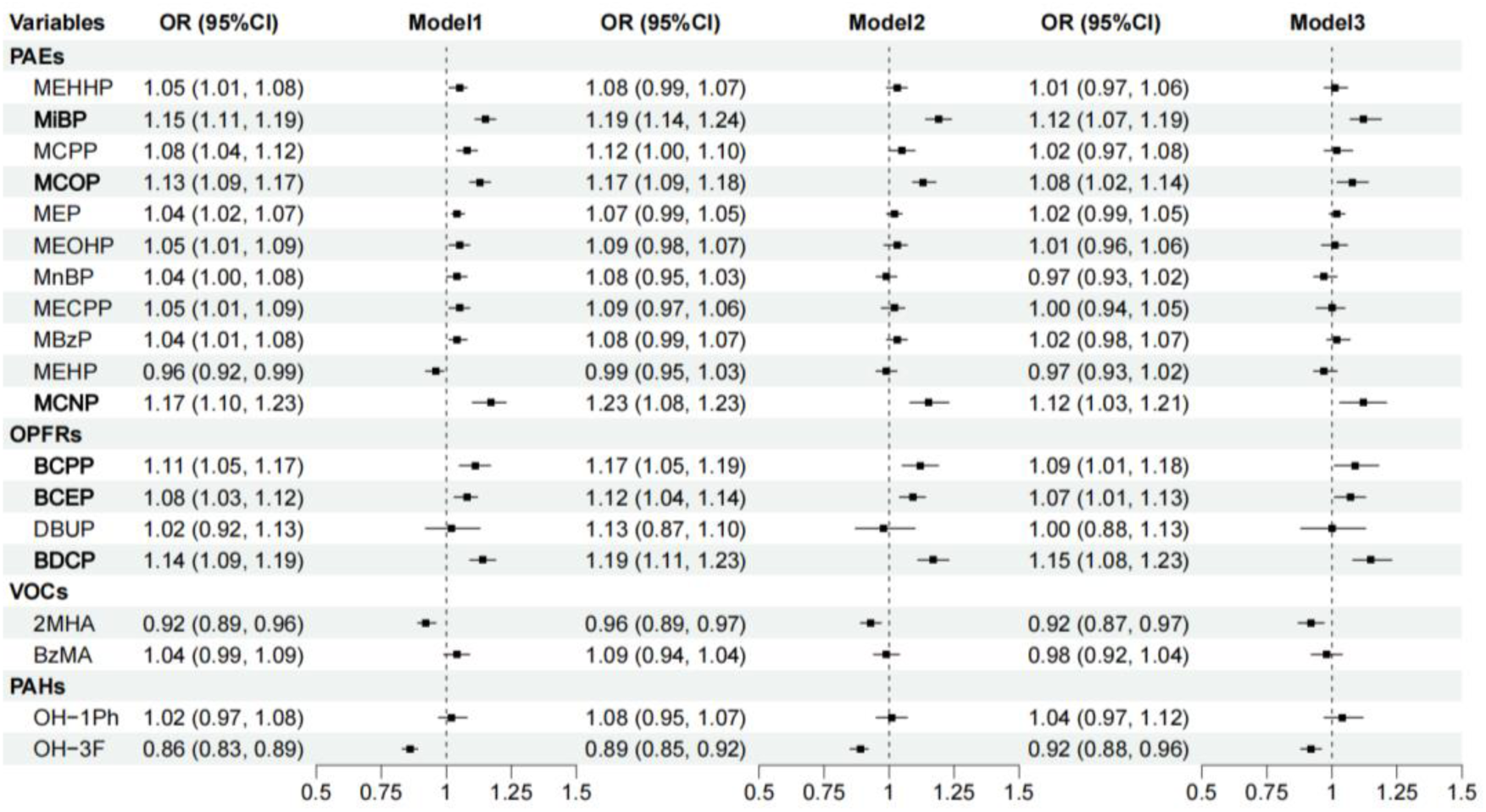
Logistic regression validation of individual chemicals for the risk of MASLD. Forest plot demonstrating the results of logistic regression for individual chemicals identified as significant in Principal Component Pursuit (PCP) analysis, assessing their association with the risk of MASLD. The dots represent the estimated odds ratios (OR), and the black horizontal lines indicate the 95% confidence intervals (CI) for each chemical. Model 1 adjusted for creatinine only. Model 2 adjusted for age, race, gender, creatinine, country of birth, education level, family income-to-poverty ratio, and marital status. Model 3 adjusted for age, race, gender, creatinine, country of birth, education level, family income-to-poverty ratio, marital status, smoking status, alcohol consumption, energy intake, and physical activity levels. Abbreviations: PAEs, phthalate esters; OPFRs, organophosphate flame retardants; VOCs, volatile organic compounds; PAHs, polycyclic aromatic hydrocarbons; MASLD, Metabolic Dysfunction-Associated Steatotic Liver Disease; OR, odds ratio; 95 % CI, 95 % confidence interval.

**Figure 5.**
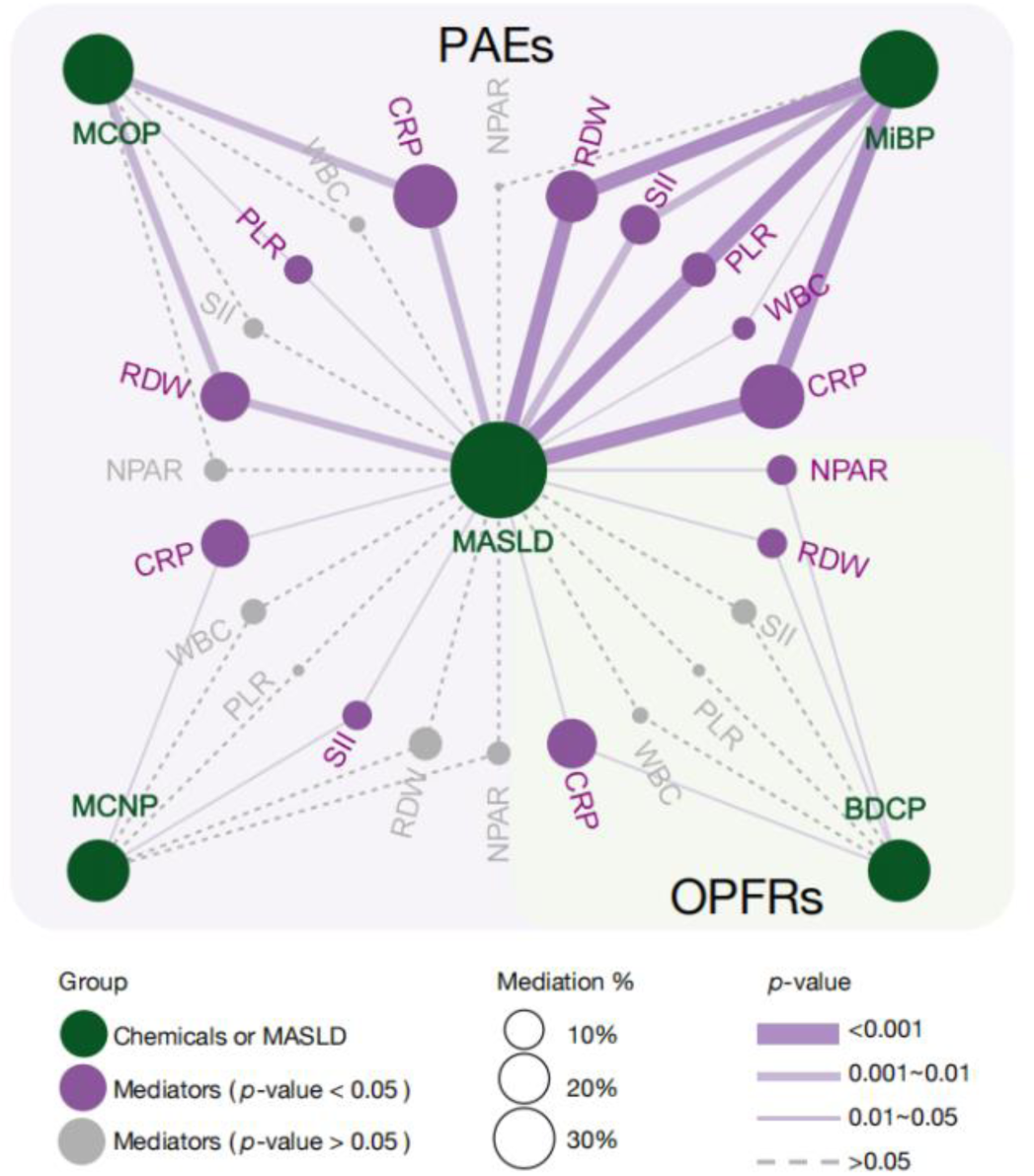
Mediation analysis of inflammatory markers in the pathway between chemical exposures and MASLD risk. Abbreviations: CRP, C-reactive protein; WBC, white blood cell count; PLR, platelet-to-lymphocyte ratio; SII , systemic immune-inflammation index; RDW, Red Cell Distribution Width; NPAR, neutrophil-percentage-to-albumin ratio; PAEs, phthalate esters; OPFRs, organophosphate flame retardants; MASLD, Metabolic Dysfunction-Associated Steatotic Liver Disease; MiBP, Mono-iso-butylphthalate; MCOP, Mono(carboxyoctyl) Phthalate; MCNP, Mono(carboxynonyl) Phthalate; BDCP, Bis (1,3-dichloro-2-propyl) Phosphate.

For PAEs, all individual chemicals within this category were included in the analysis based on the findings from the PCP. In Model 1, almost all individual PAEs chemicals were significantly associated with MASLD. This aligns with the results from the PCP analysis and the RCS findings for PAEs Dim.1, which highlighted the overall risk associated with this pollutant category. After adjusting for social and individual factors in Model 3, MiBP, MCOP, and MCNP remained significantly associated with MASLD risk. Specifically, the OR were as follows: MiBP (OR: 1.12, 95% CI: 1.07, 1.19, p<0.0001), MCOP (OR: 1.08, 95% CI: 1.02, 1.14, p=0.0048), and MCNP (OR: 1.12, 95% CI: 1.03, 1.21, p=0.0049). These findings underscore the significant role of PAEs in the risk of MASLD, with MiBP, MCOP, and MCNP showing the most notable contributions.

Similarly, for OPFRs, BDCP was positively associated with MASLD risk(OR = 1.15, 95% CI: 1.08, 1.23, p<0.0001). Positive associations were also observed for BCPP (OR = 1.09, 95% CI: 1.01, 1.18, p=0.0215) and BCEP (OR = 1.07, 95% CI: 1.01, 1.13, p=0.0137).

### 3.7 Inflammatory markers as mediators of chemical exposure and MASLD risk

Building on previous logistic regression analyses, we sought to further explore how inflammation-related factors mediate these associations. The associations between MiBP and MASLD were mediated through multiple inflammation markers. CRP mediated the largest proportion, contributing 32.42%, followed by RDW (19.62%) and SII (10.36%). The role of CRP as a mediator was also evident in the associations of MCOP, MCNP, and BDCP with MASLD. CRP mediated 31.88%, 16.52%, and 17.68% of the effects of these chemicals, respectively. Additionally, RDW mediated 17.53% of the MCOP-MASLD association and 4.78% of the BDCP-MASLD association. While PLR and WBC also exhibited mediating effects, their contributions are comparatively smaller. These findings further highlight the complex interactions between chemical exposures, inflammatory responses, and the risk of MASLD, suggesting that inflammatory markers, particularly CRP and RDW, may play a key role in the pathway through which chemical exposures contribute to the development of MASLD.

## 4 Discussion

This study systematically and comprehensively analyzed the associations between environmental chemical exposure of 60 chemicals in six categories based on NHANES databases of 1999-2020. The innovative application of PCP-PCA in analyzing chemical exposure provides a novel approach to studying the impact of multi-dimensional environmental chemicals on MASLD risk. Key findings reveal a strong association between chemical exposure, particularly PAEs and OPFRs, and MASLD. Notably, the study highlights CRP and RDW as significant mediators in the relationship between chemical exposure and MASLD risk, underscoring the importance of inflammatory markers in disease development. These results offer new insights into the mechanisms of MASLD and potential targets for intervention.

The application of PCP-PCA in this study enabled the identification of latent co-exposure patterns that are not directly characterized by traditional single-pollutant analyses. The dimension-level and individual-chemical analyses showed both convergence and divergence. For PAEs, the findings were broadly consistent: PCP-PCA identified PAEs Dim.1, which explained 90.71% of the variance, as significantly associated with MASLD, while logistic regression also identified positive associations for several individual PAE metabolites, including MiBP, MCOP, and MCNP. This concordance suggests that the dominant PAE exposure pattern may reflect contributions from multiple correlated metabolites associated with MASLD. A similar pattern was observed for OPFRs, for which significant dimension-level associations were accompanied by positive individual-chemical associations, including that of BDCP. In contrast, VOCs Dim.2 and PAHs Dim.2 were associated with MASLD at the dimension level, whereas individual chemicals within these categories showed few or no significant associations in fully adjusted models. This difference is plausible because component loadings represent the contribution of individual chemicals to a shared exposure pattern rather than their independent associations with disease risk. Dimension-reduction approaches may therefore capture shared signals from correlated exposures that are not evident in individual-chemical analyses ^[27–29]^. Conversely, PCBs and metals did not show significant dimension-level associations. Their more dispersed variance structures, with variance distributed across a larger number of dimensions than for PAEs, may partly explain the weaker associations observed for individual dimensions. Taken together, integrating PCP-PCA with individual-chemical analyses provided complementary information on both correlated exposure patterns and specific pollutants potentially related to MASLD.

Following the identification of dimension-level associations between various environmental pollutants and MASLD through PCP-PCA, we further validated these findings using two complementary approaches: RCS models to assess non-linear dose-response relationships at the dimension level, and logistic regression to identify key individual chemicals. This strategy enhanced our understanding of exposure-disease relationships and strengthened the interpretability of our results. The RCS analysis showed that PAEs Dim.1 was significantly associated with MASLD risk, with higher PCA scores generally corresponding to higher odds of MASLD. This trend is consistent with previous epidemiological studies supporting the association between high-level PAEs exposure and increased risk of liver dysfunction and NAFLD ^[41–44]^. A study involving 4,206 U.S. adults reported a significant association between urinary phthalate metabolite mixtures and NAFLD (OR=1.58, 95% CI: 1.18-2.11) ^[46]^. Similarly, a cross-sectional study based on the Korean KoNEHS found a positive correlation between increased phthalate exposure and NAFLD prevalence ^[47]^. Another study in men observed associations between PAEs exposure and elevated ALT and AST levels, suggesting potential hepatotoxicity and sex-specific vulnerability ^[48]^. In addition to PAEs Dim.1, PAEs Dim.2 and VOCs Dim.2 also exhibited upward trends in MASLD risk at higher exposure levels. Although not statistically significant, these findings may hold public health relevance.

In the analysis of individual chemicals, MiBP, MCOP, and MCNP, which are representative PAEs metabolites, were consistently associated with increased MASLD risk, possibly through disruption of lipid metabolism and induction of inflammation via PPAR pathway activation, consistent with findings from both experimental and epidemiological studies ^[45–47]^. Notably, a study based on NHANES also found a significant association between MCOP, a metabolite of diisononyl phthalate (DINP), and NAFLD (OR=4.22, 95% CI: 1.10-16.2), further supporting our findings ^[52]^. In contrast, the three major metabolites of di(2-ethylhexyl) phthalate (DEHP), namely MECPP, MEHHP, and MEOHP, were not significantly associated with MASLD after adjusting for all covariates in our analysis. Although some literature has reported positive associations ^[53]^, others have suggested that these discrepancies may stem from differences in diagnostic criteria and modeling approaches ^[48–50]^. These inconsistent findings support the need for further investigation into their potential roles in metabolic liver disease. Additionally, OPFRs such as BDCP and BCEP were also positively associated with MASLD risk in our study, in line with previous epidemiological research ^[55,56]^. Importantly, the strong consistency between the RCS and logistic regression findings reinforces the internal validity of our conclusions. While RCS models capture dose-response curves across multidimensional exposure patterns, logistic regression identifies key contributing agents. Taken together, these findings suggest that MASLD may be jointly driven by cumulative exposure patterns and specific high-risk chemicals. Our study provides an integrated analytical framework linking exposure dimensions, dose-response patterns, and individual chemical associations. These findings may help prioritize pollutants for future mechanistic studies and risk assessment in populations vulnerable to liver dysfunction.

This study identified systemic inflammation, primarily mediated by CRP and RDW, as a crucial link between environmental chemical exposure and MASLD, with CRP and RDW accounting for 32.42% and 19.62% of the observed associations, respectively. While CRP, a well-established inflammatory marker synthesized in the liver ^[38]^, has been widely recognized for its role in metabolic disorders, including hepatic inflammation and fibrosis ^[53–55]^, this study provides the first evidence that RDW mediates the association between environmental pollutant exposure and MASLD risk, addressing a critical gap in the literature. Furthermore, although RDW has been identified as a biomarker of inflammation and oxidative stress, its specific role in liver disease pathogenesis remains underexplored ^[60,61]^. Notably, elevated RDW levels have been linked to chronic liver disease severity, including advanced fibrosis and steatohepatitis ^[36,58–60]^, and have been reported to mediate 24.98% of the association between hypothyroidism and MASLD (*P*=0.012) ^[30]^, with significantly higher RDW levels observed in MAFLD patients than in controls (P<0.05) ^[37]^. The underlying mechanism may involve chronic inflammation-induced oxidative stress, which disrupts erythropoiesis and alters red blood cell turnover, thereby exacerbating liver inflammation and fibrosis ^[61]^. Chronic inflammatory responses can activate Kupffer cells and stimulate proinflammatory cytokine release, disrupting intrahepatic metabolic balance and promoting insulin resistance, hepatocyte injury, and fibrogenesis ^[31,59,65]^.

However, CRP and RDW only partially mediated the observed associations, suggesting that additional biological pathways are involved. Experimental studies have shown that PAEs and OPFRs can activate nuclear receptors such as peroxisome proliferator-activated receptors (PPARs), aryl hydrocarbon receptors (AhR), and nuclear factor kappa B (NF-κB), thereby disrupting hepatic lipid metabolism and redox homeostasis ^[24,49,66,67]^, ultimately leading to lipid accumulation and chronic inflammation ^[63–65]^. Moreover, these pollutants may impair mitochondrial function, inhibit fatty acid β-oxidation, and induce endoplasmic reticulum stress or apoptosis pathways, further contributing to hepatic steatosis and hepatocellular injury ^[66–68]^. The non-linear association observed in our study between PAEs Dim.1 and MASLD, as well as the significant effects of specific chemicals such as MiBP and BDCP, provide epidemiological support for these proposed mechanisms. Collectively, these findings suggest that the environmental etiology of MASLD involves both inflammation-dependent pathways captured by our mediation analysis and direct hepatotoxic mechanisms that warrant further investigation.

This study has several strengths, including the use of a large, nationally representative sample of 39,061 U.S. adults, the comprehensive assessment of 60 environmental chemicals across six categories, and an integrated analytical framework combining PCP-PCA, dose-response, individual-chemical, and mediation analyses to characterize both co-exposure patterns and specific chemical associations. Several limitations should also be acknowledged. First, the cross-sectional NHANES design precludes causal inference between environmental chemical exposures, inflammation-related biomarkers, and MASLD. Second, MASLD was defined using the Hepatic Steatosis Index rather than imaging or histological confirmation, which may have introduced outcome misclassification. Third, variations in detection limits, sample sizes, and analyte availability across survey cycles may have affected the precision and comparability of exposure estimates. Future prospective cohort studies with repeated exposure measurements, complemented by mechanistic investigations, are needed to clarify temporality, causal pathways, and the biological effects of key chemicals such as MiBP, MCOP, and BDCP.

## 5 Conclusion

This study utilized a novel PCP-PCA method and mediation analysis to explore the association between environmental chemical co-exposure and MASLD risk in a nationally representative NHANES cohort. Exposure to PAEs and OPFRs was significantly associated with increased MASLD risk, with the overall phthalate exposure pattern identified as the dominant contributor, showing a significant positive dose-response relationship with MASLD risk. Inflammatory markers, particularly CRP and RDW, partially mediated these associations, suggesting systemic inflammation as a potential pathway. These findings highlight the need for targeted regulation of PAEs and OPFRs and support CRP/RDW as early indicators of pollution-related liver dysfunction.

## Ethics approval and consent to participate

The NHANES protocols were approved by the National Center for Health Statistics Research Ethics Review Board, and written informed consent was obtained from all participants. The present study used publicly available de-identified data and did not require additional institutional ethical approval.

## Consent for publication

Not applicable.

## Data availability

The datasets analyzed in this study are publicly available from the National Health and Nutrition Examination Survey (NHANES) website.

## Author contributions

T.G. contributed to conceptualization, methodology, formal analysis, visualization, writing the original draft, and reviewing and editing the manuscript. Y.Z. contributed to methodology, methodological implementation, and reviewing and editing the manuscript. S.W. contributed to critical review, identification of methodological and interpretive issues, and reviewing and editing the manuscript. S.S. contributed to conceptualization, supervision, methodology, and reviewing and editing the manuscript. Q.J. contributed to methodological guidance, supervision, methodology, and reviewing and editing the manuscript. All authors read and approved the final manuscript.

## Data Availability

The datasets analyzed in this study are publicly available from the National Health and Nutrition Examination Survey (NHANES), conducted by the National Center for Health Statistics, Centers for Disease Control and Prevention.
https://wwwn.cdc.gov/nchs/nhanes/

## Reference

[1] Rinella M E, Lazarus J V, Ratziu V, et al. A multisociety delphi consensus statement on new fatty liver disease nomenclature[J]. Hepatology, 2023, 78(6): 1966.

[2] Eslam M, Sanyal A J, George J, et al. MAFLD: a consensus-driven proposed nomenclature for metabolic associated fatty liver disease[J]. Gastroenterology, 2020, 158(7): 1999–2014.e1.

[3] Eslam M, Newsome P N, Sarin S K, et al. A new definition for metabolic dysfunction-associated fatty liver disease: an international expert consensus statement[J]. Journal of Hepatology, 2020, 73(1): 202–209.

[4] Kokkorakis M, Boutari C, Katsiki N, et al. From non-alcoholic fatty liver disease (NAFLD) to steatotic liver disease (SLD): an ongoing journey towards refining the terminology for this prevalent metabolic condition and unmet clinical need[J]. Metabolism, Clinical and Experimental, 2023, 147.

[5] Polyzos S A, Mantzoros C S. Making progress in nonalcoholic fatty liver disease (NAFLD) as we are transitioning from the era of NAFLD to dys-metabolism associated fatty liver disease (DAFLD)[J]. Metabolism, 2020, 111: 154318.

[6] Chan W K, Chuah K H, Rajaram R B, et al. Metabolic dysfunction-associated steatotic liver disease (MASLD): a state-of-the-art review[J]. Journal of Obesity & Metabolic Syndrome, 2023, 32(3): 197–213.

[7] Sarin S K, Kumar M, Eslam M, et al. Liver diseases in the Asia-pacific region: a lancet gastroenterology & hepatology commission[J]. The Lancet Gastroenterology & Hepatology, 2020, 5(2): 167–228.

[8] Duell P B, Welty F K, Miller M, et al. Nonalcoholic fatty liver disease and cardiovascular risk: a scientific statement from the american heart association[J]. Arteriosclerosis, Thrombosis, and Vascular Biology, 2022, 42(6): e168–e185.

[9] Athyros V G, Polyzos S A, Kountouras J, et al. Non-alcoholic fatty liver disease treatment in patients with type 2 diabetes mellitus; new kids on the block[J]. Current Vascular Pharmacology, 2020, 18(2): 172–181.

[10] Kim K S, Hong S, Han K, et al. Association of non-alcoholic fatty liver disease with cardiovascular disease and all cause death in patients with type 2 diabetes mellitus: nationwide population based study[J]. BMJ (clinical Research Ed.), 2024, 384: e076388.

[11] Younossi Z, Anstee Q M, Marietti M, et al. Global burden of NAFLD and NASH: trends, predictions, risk factors and prevention[J]. Nature Reviews Gastroenterology & Hepatology, 2018, 15(1): 11–20.

[12] Inoue Y, Qin B, Poti J, et al. Epidemiology of obesity in adults: latest trends[J]. Current Obesity Reports, 2018, 7(4): 276–288.

[13] Polyzos S A, Kountouras J, Mantzoros C S. Obesity and nonalcoholic fatty liver disease: From pathophysiology to therapeutics[J]. Metabolism, 2019, 92: 82–97.

[14] Foulds C E, Treviño L S, York B, et al. Endocrine-disrupting chemicals and fatty liver disease[J]. Nature Reviews. Endocrinology, 2017, 13(8): 445–457.

[15] Mentsiou Nikolaou E, Kalafati I P, Dedoussis G V. The interplay between endocrine-disrupting chemicals and the epigenome towards metabolic dysfunction-associated steatotic liver disease: a comprehensive review[J]. Nutrients, 2024, 16(8): 1124.

[16] Treviño L S, Katz T A. Endocrine disruptors and developmental origins of nonalcoholic fatty liver disease[J]. Endocrinology, 2018, 159(1): 20–31.

[17] Serrano S E, Braun J, Trasande L, et al. Phthalates and diet: a review of the food monitoring and epidemiology data[J]. Environmental Health: A Global Access Science Source, 2014, 13(1): 43.

[18] Wang X, Seo Y A, Park S K. Serum selenium and non-alcoholic fatty liver disease (NAFLD) in U.S. adults: National health and nutrition examination survey (NHANES) 2011-2016[J]. Environmental Research, 2021, 197: 111190.

[19] Cheng Y, Su J, Wang X, et al. Associations between brominated flame retardants exposure and non-alcoholic fatty liver disease: mediation analysis in the NHANES[J]. Ecotoxicology and Environmental Safety, 2025, 290: 117762.

[20] Peng J, Du L L, Ma Q L. Serum glycolipids mediate the relationship of urinary bisphenols with NAFLD: analysis of a population-based, cross-sectional study[J]. Environmental Health: A Global Access Science Source, 2023, 21(1): 124.

[21] Zhang Y, Zhang M, Jiang S, et al. Associations of perfluoroalkyl substances with metabolic-associated fatty liver disease and non-alcoholic fatty liver disease: NHANES 2017-2018[J]. Cancer Causes & Control: CCC, 2024, 35(9): 1271–1282.

[22] Liu W, Cao S, Shi D, et al. Single-chemical and mixture effects of multiple volatile organic compounds exposure on liver injury and risk of non-alcoholic fatty liver disease in a representative general adult population[J]. Chemosphere, 2023, 339: 139753.

[23] Park B, Kim B, Kim C H, et al. Association between endocrine-disrupting chemical mixtures and non-alcoholic fatty liver disease with metabolic syndrome as a mediator among adults: a population-based study in korea[J]. Ecotoxicology and Environmental Safety, 2024, 276: 116310.

[24] Li W, Xiao H, Wu H, et al. Analysis of environmental chemical mixtures and nonalcoholic fatty liver disease: NHANES 1999–2014[J]. Environmental Pollution, 2022, 311: 119915.

[25] Choi Y H, Lee J Y, Moon K W. Exposure to volatile organic compounds and polycyclic aromatic hydrocarbons is associated with the risk of non-alcoholic fatty liver disease in korean adolescents: korea national environmental health survey (KoNEHS) 2015–2017[J]. Ecotoxicology and Environmental Safety, 2023, 251: 114508.

[26] Yang W, Ling X, He S, et al. PPARα/ACOX1 as a novel target for hepatic lipid metabolism disorders induced by per-and polyfluoroalkyl substances: An integrated approachPPARα/ACOX 1[J]. Environment International, 2023, 178: 108138.

[27] Wu H, Kalia V, Manz K E, et al. Exposome Profiling of Environmental Pollutants in Seminal Plasma and Novel Associations with Semen Parameters[J]. Environmental Science & Technology, 2024, 58(31): 13594–13604.

[28] Gibson E A, Zhang J, Yan J, et al. Principal Component Pursuit for Pattern Identification in Environmental Mixtures[J]. Environmental Health Perspectives, 2022, 130(11): 117008.

[29] Candès E J, Li X, Ma Y, et al. Robust principal component analysis?[J]. J. ACM, 2011, 58(3): 11:1–11:37.

[30] Wang H, Zheng C, Wang P. Exploring the nexus between hypothyroidism and metabolic dysfunction-associated steatotic liver disease: A UK biobank cohort study[J]. Scientific Reports, 2025, 15(1): 6692.

[31] Marjot T, Moolla A, Cobbold J F, et al. Nonalcoholic Fatty Liver Disease in Adults: Current Concepts in Etiology, Outcomes, and Management[J]. Endocrine Reviews, 2020, 41(1): 66–117.

[32] Low-grade systemic inflammation links heavy metal exposures to mortality: A multi-metal inflammatory index approach[J]. Science of The Total Environment, 2024, 947: 174537.

[33] Schwärzler J, Grabherr F, Grander C, et al. The pathophysiology of MASLD: An immunometabolic perspective[J]. Expert Review of Clinical Immunology, 2024, 20(4): 375–386.

[34] Chen Y, Wu M, Chen F, et al. Potential role of inflammation in relation to dietary sodium and β-carotene with non-alcoholic fatty liver disease: A mediation analysis[J]. Nutrition & Diabetes, 2022, 12(1): 1–8.

[35] Lian X, Guo J, Wang Y, et al. Association between Volatile Organic Compound Exposure and Sex Hormones in Adolescents: The Mediating Role of Serum Albumin[J]. Toxics, 2024, 12(6): 438.

[36] Giorgio V, Mosca A, Alterio A, et al. Elevated hemoglobin level is associated with advanced fibrosis in pediatric nonalcoholic fatty liver disease[J]. Journal of Pediatric Gastroenterology and Nutrition, 2017, 65(2): 150–155.

[37] Michalak A, Guz M, Kozicka J, et al. Red blood cell distribution width derivatives in alcohol-related liver cirrhosis and metabolic-associated fatty liver disease[J]. World Journal of Gastroenterology, 2022, 28(38): 5636–5647.

[38] Turco L, Garcia-Tsao G, Magnani I, et al. Cardiopulmonary hemodynamics and C-reactive protein as prognostic indicators in compensated and decompensated cirrhosis[J]. Journal of Hepatology, 2018, 68(5): 949–958.

[39] Liang B, Qiu X, Huang J, et al. Nonlinear associations of the hs-CRP/HDL-C index with metabolic dysfunction-associated steatotic liver disease and advanced liver fibrosis in US adults: insights from NHANES 2017–2018[J]. Scientific Reports, 2025, 15(1): 4029.

[40] Cdc. National health and nutrition examination survey[EB/OL]//National Health and Nutrition Examination Survey. (2025-04-30)[2025-05-27]. https://www.cdc.gov/nchs/nhanes/index.html.

[41] Hornung R W, Reed L D. Estimation of average concentration in the presence of nondetectable values[J]. Applied Occupational and Environmental Hygiene, 1990, 5(1): 46-51.

[42] Ng C H, Chan K E, Chin Y H, et al. The effect of diabetes and prediabetes on the prevalence, complications and mortality in nonalcoholic fatty liver disease[J]. Clinical and Molecular Hepatology, 2022, 28(3): 565–574.

[43] Ruhl C E, Everhart J E. Fatty liver indices in the multiethnic united states national health and nutrition examination survey[J]. Alimentary Pharmacology & Therapeutics, 2015, 41(1): 65–76.

[44] Du Z, Jiang J, Lv X, et al. The neutrophil-to-lymphocyte ratio predicts all-cause and cardiovascular mortality in patients with nonalcoholic fatty liver disease: evidence from the national health and nutrition examination survey 1999 to 2018[J]. Medicine, 2026, 105(22): e48962.

[45] Sun J, Yang S, Zhang Y, et al. Relationship between phthalates exposures and metabolic dysfunction-associated fatty liver disease in United States adults[J]. PLOS One, 2024, 19(4): e0301097.

[46] Cai S, Fan J, Ye J, et al. Phthalates exposure is associated with non-alcoholic fatty liver disease among US adults[J]. Ecotoxicology and Environmental Safety, 2021, 224: 112665.

[47] Yang Y J, Kim T, Hong Y P. Urinary phthalate levels associated with the risk of nonalcoholic fatty liver disease in adults: the korean national environmental health survey (KoNEHS) 2012-2014[J]. International Journal of Environmental Research and Public Health, 2021, 18(11): 6035.

[48] Milošević N, Milić N, Živanović Bosić D, et al. Potential influence of the phthalates on normal liver function and cardiometabolic risk in males[J]. Environmental Monitoring and Assessment, 2017, 190(1): 17.

[49] Zhang Y J, Guo J L, Xue J chuan, et al. Phthalate metabolites: characterization, toxicities, global distribution, and exposure assessment[J]. Environmental Pollution, 2021, 291: 118106.

[50] Bility M T, Thompson J T, McKee R H, et al. Activation of mouse and human peroxisome proliferator-activated receptors (PPARs) by phthalate monoesters[J]. Toxicological Sciences: an Official Journal of the Society of Toxicology, 2004, 82(1): 170–182.

[51] Hurst C H, Waxman D J. Activation of PPARalpha and PPARgamma by environmental phthalate monoesters[J]. Toxicological Sciences: an Official Journal of the Society of Toxicology, 2003, 74(2): 297–308.

[52] Zou J, Gu Q, Gu D. Association between phthalates exposure and non-alcoholic fatty liver disease under different diagnostic criteria: A cross-sectional study based on NHANES 2017 to 2018[J]. Frontiers in Public Health, 2024, 12: 1407976.

[53] Chen X, Tian F, Wu J, et al. Associations of phthalates with NAFLD and liver fibrosis: a nationally representative cross-sectional study from NHANES 2017 to 2018[J]. Frontiers in Nutrition, 2022, 9.

[54] Gogola T, Pitkänen S, Huovinen M, et al. Association between phthalate exposure and metabolic dysfunction-associated steatotic liver disease (MASLD) – systematic literature review[J]. Environmental Research, 2025, 273: 121186.

[55] Aimuzi R, Xie Z, Qu Y, et al. Associations of urinary organophosphate esters metabolites and diet quality with nonalcoholic/metabolic dysfunction-associated fatty liver diseases in adults[J]. Ecotoxicology and Environmental Safety, 2023, 254: 114720.

[56] Chai H, Hu W, Dai Y, et al. Environmental exposure to organophosphate esters and suspected non-alcoholic fatty liver disease among US adults: a mixture analysis[J]. Frontiers in Public Health, 2022, 10: 995649.

[57] Duan Y, Pan X, Luo J, et al. Association of Inflammatory Cytokines With Non-Alcoholic Fatty Liver Disease[J]. Frontiers in Immunology, 2022, 13.

[58] Rogal S S, Bielefeldt K, Wasan A D, et al. Inflammation, Psychiatric Symptoms, and Opioid Use Are Associated With Pain and Disability in Patients With Cirrhosis[J]. Clinical Gastroenterology and Hepatology, 2015, 13(5): 1009–1016.

[59] Ding J, Liu H, Zhang X, et al. Integrative multiomic analysis identifies distinct molecular subtypes of NAFLD in a Chinese population[J]. Science Translational Medicine, 2024, 16(772): eadh9940.

[60] Dallio M, Romeo M, Vaia P, et al. Red cell distribution width/platelet ratio estimates the 3-year risk of decompensation in metabolic dysfunction-associated steatotic liver disease-induced cirrhosis[J]. World Journal of Gastroenterology, 2024, 30(7): 685–704.

[61] Aslam H, Oza F, Ahmed K, et al. The role of red cell distribution width as a prognostic marker in chronic liver disease: A literature review[J]. International journal of molecular sciences, 2023, 24(4): 3487.

[62] Yang W, Huang H, Wang Y, et al. High red blood cell distribution width is closely associated with nonalcoholic fatty liver disease[J]. European Journal of Gastroenterology & Hepatology, 2014, 26(2): 174–178.

[63] Kayadibi H, Sertoglu E, Uyanik M, et al. Is increased red cell distribution width an indicating marker of nonalcoholic steatohepatitis and fibrotic stage?[J]. World Journal of Gastroenterology, 2014, 20(35): 12711–12712.

[64] Dogan S, Celikbilek M, Zararsiz G, et al. Red blood cell distribution width as a non-invasive marker for the assessment of inflammation in non-alcoholic steatohepatitis[J]. Hepato-Gastroenterology, 2015, 62(138): 393–398.

[65] Valenzuela-Vallejo L, Chrysafi P, Kouvari M, et al. Circulating hormones in biopsy-proven steatotic liver disease and steatohepatitis: a multicenter observational study[J]. Metabolism, Clinical and Experimental, 2023, 148: 155694.

[66] Chen H, Zhang W, Rui B bei, et al. Di(2-ethylhexyl) phthalate exacerbates non-alcoholic fatty liver in rats and its potential mechanisms[J]. Environmental Toxicology and Pharmacology, 2016, 42: 38–44.

[67] Zhang Y, Wang S, Zhao T, et al. Mono-2-ethylhexyl phthalate (MEHP) promoted lipid accumulation via JAK2/STAT5 and aggravated oxidative stress in BRL-3A cells[J]. Ecotoxicology and Environmental Safety, 2019, 184: 109611.

[68] Syed-Abdul M M. Lipid metabolism in metabolic-associated steatotic liver disease (MASLD)[J]. Metabolites, 2023, 14(1): 12.

[69] Kawano Y, Cohen D E. Mechanisms of hepatic triglyceride accumulation in non-alcoholic fatty liver disease[J]. Journal of Gastroenterology, 2013, 48(4): 434–441.

[70] Badmus O O, Hillhouse S A, Anderson C D, et al. Molecular mechanisms of metabolic associated fatty liver disease (MAFLD): functional analysis of lipid metabolism pathways[J]. Clinical Science (london, England: 1979), 2022, 136(18): 1347-1366.

[71] Radosavljevic T, Brankovic M, Samardzic J, et al. Altered mitochondrial function in MASLD: key features and promising therapeutic approaches[J]. Antioxidants, 2024, 13(8): 906.

[72] Zheng Y, Wang S, Wu J, et al. Mitochondrial metabolic dysfunction and non-alcoholic fatty liver disease: new insights from pathogenic mechanisms to clinically targeted therapy[J]. Journal of Translational Medicine, 2023, 21(1): 510.

[73] Kowalczyk P, Sulejczak D, Kleczkowska P, et al. Mitochondrial oxidative stress-a causative factor and therapeutic target in many diseases[J]. International journal of molecular sciences, 2021, 22(24): 13384.

