## Supplementary Materials for "Inflammation-Related Biomarkers Linking Environmental Chemical Mixtures and Metabolic Dysfunction-Associated Steatotic Liver Disease: A Population-Based Study from NHANES 1999–2020"

### Supplementary Table Contents


Table S1. Characteristics of the 60 environmental chemicals included in the analysis

Table S2. Formulas and definitions of inflammatory indices

Table S3. Variance explained by PCP-PCA-derived exposure dimensions across chemical categories

Table S4. Associations of candidate PCP-PCA-derived exposure dimensions with MASLD risk

Table S5. Component loadings of individual chemicals in selected MASLD-associated PCP-PCA dimensions

Table S6. Full mediation analysis results for inflammatory biomarkers

**Table S1. Characteristics of the 60 environmental chemicals included in the analysis**

| Categories | Abbreviation | Fullname | LOD | GM | Min | 25th | 50th | 75th | Max |
|---|---|---|---|---|---|---|---|---|---|
| PAHs,ng/L | | | | | | | | | |
| | OH-1N | 1-Hydroxynaphthalene (1-Naphthol) | 60.00 | 2188.95 | 4.20 | 762.00 | 1832.20 | 6311.00 | 161363.00 |
| | OH-2N | 2-Hydroxynaphthalene (2-Naphthol) | 90.00 | 4246.89 | 9.30 | 1813.10 | 4234.50 | 10216.20 | 128300.00 |
| | OH-3F | 3-Hydroxyfluorene | 8.00 | 111.19 | 1.40 | 42.00 | 88.30 | 268.00 | 4803.00 |
| | OH-2F | 2-Hydroxyfluorene | 8.00 | 272.57 | 2.80 | 116.00 | 239.00 | 608.00 | 8261.70 |
| | OH-1Ph | 1-Hydroxyphenanthrene | 9.00 | 126.74 | 2.80 | 68.00 | 129.00 | 239.50 | 2112.30 |
| | OH-1P | 1-Hydroxypyrene | 70.00 | 100.31 | 2.10 | 49.50 | 98.00 | 206.00 | 2730.00 |
| | OH-2,3Ph | 2-Hydroxyphenanthrene & 3-Hydroxyphenanthrene | 10.00 | 121.49 | 7.10 | 64.00 | 121.50 | 221.20 | 2010.00 |
| OPFRs,μg/L | | | | | | | | | |
| | BCPP | Bis (1-chloro-2-propyl) Phosphate | 0.10 | 0.15 | 0.07 | 0.07 | 0.07 | 0.27 | 3.28 |
| | BCEP | Bis-2-chloroethyl Phosphate | 0.10 | 0.35 | 0.06 | 0.13 | 0.34 | 0.80 | 18.70 |
| | BDCP | Bis (1,3-dichloro-2-propyl) Phosphate | 0.10 | 0.85 | 0.07 | 0.36 | 0.86 | 1.97 | 50.70 |
| | DBUP | Dibutyl Phosphate | 0.10 | 0.13 | 0.07 | 0.07 | 0.07 | 0.18 | 3.39 |
| | DPHP | Diphenyl Phosphate | 0.10 | 0.75 | 0.07 | 0.35 | 0.74 | 1.56 | 38.70 |
| PCB,ng/g | | | | | | | | | |
| | PCB99 | 2,2’,4,4’,5-Pentachlorobiphenyl | 0.18 | 0.04 | 0.00 | 0.02 | 0.03 | 0.06 | 0.34 |
| | PCB138 | 2,2’,3,4,4’,5’- and 2,3,3’,4,4’,6-Hexachlorobiphenyl | 0.18 | 0.14 | 0.01 | 0.08 | 0.13 | 0.27 | 2.13 |
| | PCB153 | 2,2’,4,4’,5,5’-Hexachlorobiphenyl | 0.18 | 0.20 | 0.01 | 0.10 | 0.20 | 0.40 | 3.71 |
| | PCB170 | 2,2’,3,3’,4,4’,5-Heptachlorobiphenyl | 0.18 | 0.06 | 0.00 | 0.03 | 0.06 | 0.12 | 0.88 |
| | PCB180 | 2,2’,3,4,4’,5,5’-Heptachlorobipheny | 0.18 | 0.14 | 0.00 | 0.06 | 0.16 | 0.32 | 3.65 |
| | PCB187 | 2,2’,3,4’,5,5’,6-Heptachlorobiphenyl | 1.18 | 0.05 | 0.00 | 0.02 | 0.05 | 0.09 | 0.76 |
| | PCB74 | 2,4,4’,5-Tetrachlorobiphenyl | 0.44 | 0.05 | 0.00 | 0.02 | 0.04 | 0.09 | 0.57 |
| | PCB188 | 2,3’,4,4’,5-Pentachlorobiphenyl | 0.19 | 0.05 | 0.00 | 0.02 | 0.05 | 0.11 | 0.78 |

| Categories | Abbreviation | Fullname | LOD | GM | Min | 25th | 50th | 75th | Max |
|---|---|---|---|---|---|---|---|---|---|
| PAEs,μg/L | | | | | | | | | |
| | MCNP | Mono(carboxynonyl) Phthalate | 0.20 | 2.27 | 0.14 | 1.10 | 2.20 | 4.50 | 64.35 |
| | MCOP | Mono(carboxyoctyl) Phthalat | 0.30 | 9.97 | 0.14 | 3.90 | 8.80 | 24.60 | 674.02 |
| | MCPP | Mono-(3-carboxypropyl) phthalate | 0.40 | 2.10 | 0.10 | 1.00 | 2.16 | 4.50 | 95.70 |
| | MEP | Mono-ethyl phthalate | 1.20 | 85.04 | 0.37 | 27.10 | 79.50 | 249.41 | 9040.70 |
| | MiBP | Mono-iso-butylphthalate | 0.80 | 5.94 | 0.14 | 2.90 | 6.50 | 13.40 | 240.70 |
| | MnBP | Mono-n-butyl phthalate | 0.40 | 14.48 | 0.28 | 7.30 | 15.90 | 32.00 | 669.50 |
| | MBzP | Mono-benzyl phthalate | 0.30 | 6.10 | 0.06 | 2.60 | 6.48 | 14.99 | 435.67 |
| | MEHP | Mono-(2-ethyl)-hexylphthalate | 0.80 | 2.04 | 0.35 | 0.78 | 1.80 | 4.41 | 94.30 |
| | MEHHP | Mono-(2-ethyl-5-hydroxyhexyl) phthalate | 0.40 | 12.22 | 0.14 | 5.40 | 12.10 | 26.93 | 788.20 |
| | MEOHP | Mono-(2-ethyl-5-oxohexyl) phthalate | 0.20 | 7.63 | 0.14 | 3.40 | 7.60 | 16.70 | 471.30 |
| | MECPP | Mono-(2-ethyl-5-carboxy-pentyl) phthalate | 0.40 | 18.20 | 0.14 | 8.40 | 17.60 | 37.89 | 885.20 |
| VOCs,μg/L | | | | | | | | | |
| | MUCA | trans, trans-Muconic acid | 9.81 | 55.71 | 6.94 | 24.20 | 53.00 | 119.00 | 2610.00 |
| | 3HPMA | N-Acetyl-S-(3-hydroxypropyl)-L-cysteine | 13.00 | 265.22 | 9.20 | 126.00 | 254.00 | 528.00 | 6300.00 |
| | MADA | Mandelic acid | 12.00 | 132.59 | 8.50 | 75.10 | 140.00 | 239.20 | 1840.00 |
| | BzMA | N-Acetyl-S-(benzyl)-L-cysteine | 0.50 | 6.72 | 0.35 | 3.43 | 6.69 | 12.70 | 157.00 |
| | 3MHA+4MHA | 3-Methylhippuric acid + 4-Methylhippuric acid | 8.00 | 188.97 | 5.66 | 77.60 | 179.00 | 470.00 | 7090.00 |
| | 2MHA | 2-Methylhippuric acid | 5.00 | 30.05 | 3.54 | 12.70 | 28.60 | 72.40 | 1020.00 |
| | 4HBeMA | N-Acetyl-s-(4-hydroxy-2-butenyl)-Lcysteine | 0.60 | 5.77 | 0.42 | 2.58 | 5.17 | 11.60 | 175.00 |
| Metals,μg/L | | | | | | | | | |
| | bCo | Cobalt | 0.06 | 0.14 | 0.04 | 0.11 | 0.14 | 0.18 | 0.84 |
| | bPb | Lead | 0.07 | 1.30 | 0.05 | 0.80 | 1.30 | 2.10 | 11.05 |
| | bCd | Cadmium | 0.10 | 0.36 | 0.07 | 0.20 | 0.35 | 0.60 | 3.14 |
| | bHg | Mercury | 0.28 | 0.86 | 0.07 | 0.42 | 0.81 | 1.66 | 13.31 |
| | bSe | Selenium | 24.48 | 188.24 | 89.80 | 173.30 | 188.40 | 204.50 | 298.00 |

| Categories | Abbreviation | Fullname | LOD | GM | Min | 25th | 50th | 75th | Max |
|---|---|---|---|---|---|---|---|---|---|
| | bMn | Manganese | 0.99 | 9.31 | 1.57 | 7.42 | 9.25 | 11.62 | 26.98 |
| | uAs | Arsenic | 0.23 | 8.19 | 0.16 | 3.83 | 7.64 | 16.22 | 228.50 |
| | uCr | Chromium | 0.19 | 0.19 | 0.13 | 0.13 | 0.13 | 0.26 | 1.62 |
| | uHg | Mercury | 0.13 | 0.40 | 0.04 | 0.16 | 0.38 | 0.88 | 11.21 |
| | uBa | Barium | 0.06 | 1.12 | 0.04 | 0.56 | 1.16 | 2.25 | 27.80 |
| | uCd | Cadmium | 0.04 | 0.24 | 0.03 | 0.12 | 0.24 | 0.49 | 4.36 |
| | uCo | Cobalt | 0.02 | 0.33 | 0.02 | 0.20 | 0.35 | 0.56 | 4.70 |
| | uCs | Cesium | 0.09 | 4.21 | 0.07 | 2.76 | 4.60 | 6.86 | 33.50 |
| | uMo | Molybdenum | 0.80 | 38.42 | 0.57 | 21.83 | 41.79 | 71.98 | 593.00 |
| | uPb | Lead | 0.03 | 0.48 | 0.02 | 0.25 | 0.50 | 0.90 | 8.50 |
| | uSb | Antimony | 0.02 | 0.06 | 0.02 | 0.03 | 0.06 | 0.11 | 0.94 |
| | uSn | Tin | 0.09 | 0.51 | 0.06 | 0.22 | 0.47 | 1.02 | 23.83 |
| | uTl | Thallium | 0.02 | 0.15 | 0.01 | 0.10 | 0.16 | 0.26 | 1.37 |
| | uW | Tungsten | 0.02 | 0.07 | 0.01 | 0.03 | 0.07 | 0.13 | 1.97 |
| | uNi | Nickel | 0.31 | 1.11 | 0.22 | 0.65 | 1.16 | 1.92 | 14.00 |
| | uSr | Strontium | 2.34 | 82.38 | 1.66 | 47.93 | 88.50 | 154.84 | 1007.81 |
| | uU | Uranium | 0.00 | 0.01 | 0.00 | 0.00 | 0.01 | 0.01 | 0.13 |

**Note:** Chemicals were excluded from the analysis if their detection rate <50%. Abbreviations: GM, geometric mean.

**Table S2. Formulas and definitions of inflammatory indices**

| Abbreviation | Full Name | Unit | Calculation Formula |
|---|---|---|---|
| GGT | Gamma-glutamyl transferase | IU/L | Directly measured |
| CRP | C-reactive protein | mg/L | Directly measured |
| WBC | White blood cell count | $10^3$ cells/μL | Directly measured |
| SIRI | Systemic Inflammation Response Index | dimensionless | Neutrophil count × Monocyte count / Lymphocyte count |
| PLR | Platelet-to-lymphocyte ratio | dimensionless | Platelet count / Lymphocyte count |
| NLR | Neutrophil-to-lymphocyte ratio | dimensionless | Neutrophil count / Lymphocyte count |
| dNLR | Derived neutrophil-to-lymphocyte ratio | dimensionless | Neutrophil count / (WBC count − Neutrophil count) |
| SII | Systemic immune-inflammation index | dimensionless | Platelet count × Neutrophil count / Lymphocyte count |
| MLR | Monocyte-to-lymphocyte ratio | dimensionless | Monocyte count / Lymphocyte count |
| NMLR | Neutrophil-monocyte to lymphocyte ratio | dimensionless | (Neutrophil count + Monocyte count) / Lymphocyte count |
| AISI | Aggregate index of systemic inflammation | dimensionless | Neutrophil count × Monocyte count × Platelet count / Lymphocyte count |
| ELR | Eosinophil-to-lymphocyte ratio | dimensionless | Eosinophil count / Lymphocyte count |
| NPAR | Neutrophil-percentage-to-albumin ratio | % / (g/dL) | Neutrophil percentage (%) × 100 / Serum albumin (g/dL) |
| RDW | Red Cell Distribution Width | % | Directly measured (reported as CV%) |
| RPR | Red Cell Distribution Width to Platelet Count Ratio | % / ($10^9$/L) | RDW (%) / Platelet count |

**Table S3. Variance explained by PCP-PCA-derived exposure dimensions across chemical categories**

| Chemical category | Dimension | Eigenvalue | Variance explained, % | Cumulative variance, % |
|---|---|---|---|---|
| PAEs | Dim.1 | 9.978 | 90.713 | 90.713 |
| | Dim.2 | 0.524 | 4.761 | 95.474 |
| | Dim.3 | 0.425 | 3.861 | 99.335 |
| | Dim.4 | 0.066 | 0.598 | 99.932 |
| | Dim.5 | 0.007 | 0.067 | 100.000 |
| | Dim.6 | <0.001 | <0.001 | 100.000 |
| | Dim.7 | <0.001 | <0.001 | 100.000 |
| | Dim.8 | <0.001 | <0.001 | 100.000 |
| | Dim.9 | <0.001 | <0.001 | 100.000 |
| | Dim.10 | <0.001 | <0.001 | 100.000 |
| | Dim.11 | <0.001 | <0.001 | 100.000 |
| OPFRs | Dim.1 | 1.721 | 34.415 | 34.415 |
| | Dim.2 | 1.001 | 20.021 | 54.436 |
| | Dim.3 | 0.880 | 17.592 | 72.029 |
| | Dim.4 | 0.724 | 14.472 | 86.500 |
| | Dim.5 | 0.675 | 13.500 | 100.000 |
| VOCs | Dim.1 | 6.824 | 97.491 | 97.491 |
| | Dim.2 | 0.150 | 2.148 | 99.639 |
| | Dim.3 | 0.025 | 0.360 | 100.000 |
| | Dim.4 | <0.001 | <0.001 | 100.000 |
| | Dim.5 | <0.001 | <0.001 | 100.000 |
| | Dim.6 | <0.001 | <0.001 | 100.000 |
| | Dim.7 | <0.001 | <0.001 | 100.000 |
| PAHs | Dim.1 | 6.921 | 98.876 | 98.876 |
| | Dim.2 | 0.057 | 0.818 | 99.694 |
| | Dim.3 | 0.021 | 0.306 | 100.000 |
| | Dim.4 | <0.001 | <0.001 | 100.000 |
| | Dim.5 | <0.001 | <0.001 | 100.000 |
| | Dim.6 | <0.001 | <0.001 | 100.000 |
| | Dim.7 | <0.001 | <0.001 | 100.000 |
| PCBs | Dim.1 | 2.149 | 30.703 | 30.703 |
| | Dim.2 | 1.994 | 28.486 | 59.189 |
| | Dim.3 | 1.082 | 15.452 | 74.641 |
| | Dim.4 | 0.984 | 14.051 | 88.691 |
| | Dim.5 | 0.656 | 9.374 | 98.066 |
| | Dim.6 | 0.126 | 1.807 | 99.872 |
| | Dim.7 | 0.009 | 0.128 | 100.000 |
| | Dim.8 | <0.001 | <0.001 | 100.000 |
| Metals | Dim.1 | 10.936 | 64.327 | 64.327 |
| | Dim.2 | 1.287 | 7.570 | 71.897 |
| | Dim.3 | 1.010 | 5.940 | 77.837 |

| Chemical category | Dimension | Eigenvalue | Variance explained, % | Cumulative variance, % |
|---|---|---|---|---|
| | Dim.4 | 0.975 | 5.735 | 83.572 |
| | Dim.5 | 0.915 | 5.381 | 88.954 |
| | Dim.6 | 0.780 | 4.587 | 93.541 |
| | Dim.7 | 0.674 | 3.967 | 97.508 |
| | Dim.8 | 0.244 | 1.435 | 98.943 |
| | Dim.9 | 0.126 | 0.740 | 99.683 |
| | Dim.10 | 0.047 | 0.279 | 99.963 |
| | Dim.11 | 0.004 | 0.026 | 99.989 |
| | Dim.12 | 0.002 | 0.011 | 100.000 |
| | Dim.13 | <0.001 | <0.001 | 100.000 |
| | Dim.14 | <0.001 | <0.001 | 100.000 |
| | Dim.15 | <0.001 | <0.001 | 100.000 |
| | Dim.16 | <0.001 | <0.001 | 100.000 |
| | Dim.17 | <0.001 | <0.001 | 100.000 |

Notes: Eigenvalues and variance percentages were derived from PCA performed after principal component pursuit decomposition within each chemical category. Variance explained indicates the proportion of category-specific exposure variation explained by each dimension. Cumulative variance indicates the total proportion of exposure variation explained up to the corresponding dimension. Abbreviations: Dim., dimension; PCP-PCA, principal component pursuit-principal component analysis; PCA, principal component analysis; PAEs, phthalate esters; OPFRs, organophosphate flame retardants; VOCs, volatile organic compounds; PAHs, polycyclic aromatic hydrocarbons; PCBs, polychlorinated biphenyls.

**Table S4. Associations between the first six PCP-PCA dimensions and MASLD risk**

| Chemical category | Dimension | Estimate | Standard error | *P*-value | *FDR*-adjusted *P*-value | *FDR* significance | Selected for further analysis |
|---|---|---|---|---|---|---|---|
| **PAEs** | **Dim.1** | 1.057 | 0.009 | <0.001 | <0.001 | *** | Yes |
| | **Dim.2** | 1.149 | 0.040 | <0.001 | 0.005 | ** | Yes |
| | Dim.3 | 0.957 | 0.045 | 0.326 | 0.518 | NS | No |
| | Dim.4 | 0.692 | 0.114 | 0.001 | 0.007 | ** | No |
| | Dim.5 | 0.754 | 0.340 | 0.406 | 0.618 | NS | No |
| | Dim.6 | 3285.398 | 10.369 | 0.435 | 0.634 | NS | No |
| **OPFRs** | **Dim.1** | 0.870 | 0.033 | <0.001 | <0.001 | *** | Yes |
| | Dim.2 | 2.101 | 0.481 | 0.123 | 0.287 | NS | No |
| | **Dim.3** | 0.880 | 0.047 | 0.007 | 0.030 | * | Yes |
| | Dim.4 | 1.001 | 0.051 | 0.987 | 0.987 | NS | No |
| | Dim.5 | 1.086 | 0.054 | 0.122 | 0.306 | NS | No |
| **VOCs** | Dim.1 | 1.008 | 0.019 | 0.675 | 0.762 | NS | No |
| | **Dim.2** | 1.490 | 0.121 | <0.001 | 0.007 | ** | Yes |
| | Dim.3 | 0.667 | 0.293 | 0.167 | 0.325 | NS | No |
| | Dim.4 | 433.428 | 11.884 | 0.609 | 0.790 | NS | No |
| | Dim.5 | NE | NE | NE | NE | NE | No |
| | Dim.6 | NE | NE | NE | NE | NE | No |
| **PAHs** | Dim.1 | 0.996 | 0.016 | 0.819 | 0.868 | NS | No |
| | **Dim.2** | 0.449 | 0.167 | <0.001 | <0.001 | *** | Yes |
| | Dim.3 | 0.891 | 0.272 | 0.673 | 0.785 | NS | No |
| | Dim.4 | NE | NE | NE | NE | NE | No |
| | Dim.5 | NE | NE | NE | NE | NE | No |
| | Dim.6 | NE | NE | NE | NE | NE | No |
| PCBs | Dim.1 | 1.026 | 0.018 | 0.160 | 0.350 | NS | No |
| | Dim.2 | 1.071 | 0.037 | 0.063 | 0.202 | NS | No |
| | Dim.3 | 0.927 | 0.064 | 0.232 | 0.406 | NS | No |
| | Dim.4 | 0.975 | 0.075 | 0.732 | 0.801 | NS | No |
| | Dim.5 | 1.685 | 0.237 | 0.028 | 0.109 | NS | No |
| | Dim.6 | 0.914 | 0.414 | 0.827 | 0.851 | NS | No |
| Metals | Dim.1 | 1.005 | 0.012 | 0.649 | 0.811 | NS | No |
| | Dim.2 | 0.939 | 0.032 | 0.052 | 0.183 | NS | No |
| | Dim.3 | 1.050 | 0.035 | 0.164 | 0.337 | NS | No |
| | Dim.4 | 0.953 | 0.036 | 0.187 | 0.345 | NS | No |
| | Dim.5 | 0.959 | 0.039 | 0.273 | 0.455 | NS | No |
| | Dim.6 | 0.934 | 0.040 | 0.090 | 0.241 | NS | No |

Notes: Estimates represent the model-based association estimates for each PCP-PCA dimension score in relation to MASLD status. Standard errors refer to the corresponding model-based standard errors. Candidate dimensions comprised the first six dimensions within each chemical category when available; categories with fewer than six measurable dimensions contributed all available dimensions. NE indicates that the estimate, standard error, P-value, or FDR-adjusted P-value was not estimable due to numerical instability. P-values were derived from regression models evaluating the association between each PCP-PCA dimension score and MASLD status. P-values were adjusted using the Benjamini-Hochberg false discovery rate method. FDR significance was defined as follows: ***FDR-adjusted P<0.001, **FDR-adjusted P<0.01, *FDR-adjusted P<0.05, and NS, not significant. Dimensions marked as selected were retained for downstream dose-response and loading analyses based on statistical evidence, variance contribution, and numerical stability. Abbreviations: Dim., dimension; MASLD, metabolic dysfunction-associated steatotic liver disease; PCP-PCA, principal component pursuit-principal component analysis; FDR, false discovery rate; PAEs, phthalate esters; OPFRs, organophosphate flame retardants; VOCs, volatile organic compounds; PAHs, polycyclic aromatic hydrocarbons; PCBs, polychlorinated biphenyls; NE, not estimable due to numerical instability; NS, not significant.

**Table S5. Component loadings of individual chemicals in selected MASLD-associated PCP-PCA dimensions**

| Chemical category | Dimension | Chemical abbreviation | Loading | Absolute loading | Loading direction |
|---|---|---|---|---|---|
| PAEs | Dim.1 | MECPP | 0.983 | 0.983 | Positive |
| | | MBzP | 0.975 | 0.975 | Positive |
| | | MEP | 0.974 | 0.974 | Positive |
| | | MEHHP | 0.967 | 0.967 | Positive |
| | | MnBP | 0.966 | 0.966 | Positive |
| | | MiBP | 0.958 | 0.958 | Positive |
| | | MEOHP | 0.957 | 0.957 | Positive |
| | | MCOP | 0.950 | 0.950 | Positive |
| | | MEHP | 0.923 | 0.923 | Positive |
| | | MCPP | 0.916 | 0.916 | Positive |
| | | MCNP | 0.904 | 0.904 | Positive |
| PAEs | Dim.2 | MCNP | 0.421 | 0.421 | Positive |
| | | MCPP | 0.365 | 0.365 | Positive |
| | | MCOP | 0.253 | 0.253 | Positive |
| | | MEOHP | -0.198 | 0.198 | Negative |
| | | MEHHP | -0.194 | 0.194 | Negative |
| | | MECPP | -0.138 | 0.138 | Negative |
| | | MnBP | -0.137 | 0.137 | Negative |
| | | MEP | -0.114 | 0.114 | Negative |
| | | MBzP | -0.104 | 0.104 | Negative |
| | | MEHP | -0.102 | 0.102 | Negative |
| | | MiBP | -0.004 | 0.004 | Negative |
| OPFRs | Dim.1 | BCPP | 0.705 | 0.705 | Positive |
| | | BCEP | 0.689 | 0.689 | Positive |
| | | DBUP | 0.675 | 0.675 | Positive |
| | | BDCP | 0.541 | 0.541 | Positive |
| | | DPHP | -0.021 | 0.021 | Negative |
| OPFRs | Dim.3 | BDCP | 0.776 | 0.776 | Positive |
| | | DBUP | -0.437 | 0.437 | Negative |
| | | BCPP | -0.266 | 0.266 | Negative |
| | | BCEP | 0.093 | 0.093 | Positive |
| | | DPHP | 0.086 | 0.086 | Positive |
| VOCs | Dim.2 | BzMA | 0.237 | 0.237 | Positive |
| | | 2MHA | -0.182 | 0.182 | Negative |
| | | MADA | 0.140 | 0.140 | Positive |
| | | 4HBeMA | -0.083 | 0.083 | Negative |
| | | 3MHA+4MHA | -0.075 | 0.075 | Negative |
| | | 3HPMA | 0.037 | 0.037 | Positive |
| | | MUCA | 0.029 | 0.029 | Positive |

| Chemical category | Dimension | Chemical abbreviation | Loading | Absolute loading | Loading direction |
|---|---|---|---|---|---|
| PAHs | Dim.2 | OH-3F | 0.145 | 0.145 | Positive |
| | | OH-1Ph | -0.14 | 0.140 | Negative |
| | | OH-2F | 0.087 | 0.087 | Positive |
| | | OH-2,3Ph | -0.072 | 0.072 | Negative |
| | | OH-1P | -0.042 | 0.042 | Negative |
| | | OH-1N | 0.041 | 0.041 | Positive |
| | | OH-2N | -0.019 | 0.019 | Negative |

Notes: Loadings quantify the contribution of each individual chemical to the selected MASLD-associated PCP-PCA dimension. Absolute loading represents the magnitude of the loading irrespective of sign, and loading direction indicates whether the chemical contributes positively or negatively to the corresponding dimension score. Larger absolute values indicate stronger contribution to the latent exposure pattern. Loading direction should not be interpreted as the direction of association with MASLD by itself. Chemical abbreviations are defined in Table S1. Abbreviations: Dim., dimension; MASLD, metabolic dysfunction-associated steatotic liver disease; PCP-PCA, principal component pursuit-principal component analysis; PAEs, phthalate esters; OPFRs, organophosphate flame retardants; VOCs, volatile organic compounds; PAHs, polycyclic aromatic hydrocarbons.

**Table S6. Full mediation analysis results for inflammatory biomarkers**

| Chemical | Mediator | Estimate | Standard error | *Z*-value | *P*-value | Lower 95% CI | Upper 95% CI |
|---|---|---|---|---|---|---|---|
| **MiBP** | **CRP** | **32.42%** | 0.071 | 4.552 | **< 0.001** | 18.46% | 46.38% |
| | **WBC** | **2.38%** | 0.011 | 2.098 | **0.036** | 0.16% | 4.60% |
| | **PLR** | **7.14%** | 0.017 | 4.182 | **< 0.001** | 3.79% | 10.48% |
| | **SII** | **10.36%** | 0.035 | 2.937 | **0.003** | 3.45% | 17.28% |
| | RDW | 19.62% | 0.047 | 4.22 | **< 0.001** | 10.51% | 28.74% |
| | NPAR | 0.03% | 0.004 | 0.072 | 0.943 | -0.66% | 0.71% |
| **MCOP** | **CRP** | **31.88%** | 0.119 | 2.677 | **0.007** | 8.54% | 55.21% |
| | WBC | -0.76% | 0.014 | -0.555 | 0.579 | -3.43% | 1.92% |
| | **PLR** | **4.40%** | 0.018 | 2.448 | **0.014** | 0.88% | 7.93% |
| | SII | 1.63% | 0.012 | 1.411 | 0.158 | -0.63% | 3.89% |
| | **RDW** | **17.53%** | 0.065 | 2.701 | **0.007** | 4.81% | 30.24% |
| | NPAR | 2.35% | 0.017 | 1.393 | 0.164 | -0.96% | 5.65% |
| **MCNP** | **CRP** | **16.52%** | 0.075 | 2.195 | **0.028** | 1.77% | 31.27% |
| | WBC | -3.13% | 0.019 | -1.633 | 0.102 | -6.88% | 0.63% |
| | PLR | -0.18% | 0.014 | -0.125 | 0.9 | -2.91% | 2.56% |
| | **SII** | **4.68%** | 0.021 | 2.198 | **0.028** | 0.51% | 8.86% |
| | RDW | 6.48% | 0.034 | 1.881 | 0.06 | -0.27% | 13.23% |
| | NPAR | 2.53% | 0.017 | 1.448 | 0.148 | -0.90% | 5.96% |
| **BDCP** | **CRP** | **17.68%** | 0.09 | 1.968 | **0.049** | 0.08% | 35.29% |
| | WBC | 0.72% | 0.017 | 0.434 | 0.664 | -2.53% | 3.96% |
| | PLR | -0.24% | 0.008 | -0.315 | 0.753 | -1.72% | 1.25% |
| | SII | 2.99% | 0.022 | 1.331 | 0.183 | -1.41% | 7.39% |
| | **RDW** | **4.78%** | 0.021 | 2.223 | **0.026** | 0.56% | 8.99% |
| | **NPAR** | **4.87%** | 0.022 | 2.236 | **0.025** | 0.60% | 9.14% |

Abbreviations: CRP, C-reactive protein; WBC, white blood cell count; PLR, platelet-to-lymphocyte ratio; SII, systemic inflammatory index; NPAR, neutrophil-percentage-to-albumin ratio; RDW, Red Cell Distribution Width.
